# Utilising Nanopore direct RNA sequencing of blood from patients with sepsis for discovery of co- and post-transcriptional disease biomarkers

**DOI:** 10.1101/2024.12.13.24318230

**Authors:** Jingni He, Devika Ganesamoorthy, Jessie J-Y Chang, Josh Zhang, Sharon L Trevor, Kristen S Gibbons, Stephen J McPherson, Jessica C. Kling, Luregn J Schlapbach, Antje Blumenthal, the RAPIDS Study Group, Lachlan JM Coin

**Affiliations:** Department of Clinical Pathology, The University of Melbourne, Parkville, Australia; Department of Neuroscience, School of Translational Medicine, Monash University, Melbourne, Australia; Institute for Molecular Bioscience, The University of Queensland, Brisbane, Australia; Children’s Intensive Care Research Program, Child Health Research Centre, The University of Queensland, Brisbane, Australia; Department of Microbiology and Immunology, The University of Melbourne, Parkville, Australia; Frazer Institute, The University of Queensland, Brisbane, Australia; Department of Intensive Care and Neonatology, and Children’s Research Center, University Children’s Hospital Zurich, University of Zurich, Zurich, Switzerland; Department of Infectious Disease, Imperial College London, London United Kingdom

**Keywords:** Direct RNA-sequencing, Oxford Nanopore Technologies, expression correlation, polyadenylation, long-read sequencing, differential transcript usage, novel isoform detection

## Abstract

**Background:** RNA sequencing of whole blood has been increasingly employed to find transcriptomic signatures of disease states. These studies traditionally utilize short-read sequencing of cDNA, missing important aspects of RNA expression such as differential isoform abundance and poly(A) tail length variation.

**Methods:** We used Oxford Nanopore Technologies long-read sequencing to sequence native mRNA extracted from whole blood from 12 patients with suspected bacterial and viral sepsis, and compared with results from matching Illumina short-read cDNA sequencing data. Additionally, we explored poly(A) tail length variation, novel transcript identification and differential transcript usage.

**Results:** The correlation of gene count data between Illumina cDNA and Nanopore RNA-sequencing strongly depended on the choice of analysis pipeline; *NanoCount* for Nanopore and *Kallisto* for Illumina data yielded the highest mean Pearson’s correlation of 0.93 at gene level and 0.74 at transcript isoform level. We identified 18 genes significantly differentially polyadenylated and 4 genes with significant differential transcript usage between bacterial and viral infection. Gene ontology gene set enrichment analysis of poly(A) tail length revealed enrichment of long tails in signal transduction and short tails in oxidoreductase molecular functions. Additionally, we detected 594 non-artifactual novel transcript isoforms, including 9 novel isoforms for Immunoglobulin lambda like polypeptide 5 (*IGLL5)*.

**Conclusions:** Nanopore RNA- and Illumina cDNA-gene counts are strongly correlated, indicating that both platforms are suitable for discovery and validation of gene count biomarkers. Nanopore direct RNA-seq provides additional advantages by uncovering additional post- and co-transcriptional biomarkers, such as poly(A) tail length variation and transcript isoform usage.

## BACKGROUND

Transcriptomics provides a time and cost-effective method of understanding disease status of the patient and enables an avenue to develop targeted prophylactic, diagnostic and therapeutic strategies. Studies investigating host transcriptional response typically employ high-throughput short-read sequencing, such as Illumina sequencing, to identify gene-count biomarkers of disease (1-5). These platforms provide highly accurate sequence data with high coverage (6). However, short-read approaches rely on converting to complementary DNA (cDNA) followed by cDNA amplification using polymerase chain reaction (PCR), both of which may introduce biases that interfere with the accurate quantification of transcripts (7). Moreover, short-read sequencing has transcript/gene length-dependent expression bias (8), as well as complex compositional biases such as with guanine-cytosine (GC) content (9). Additionally, the short read lengths limit the resolution of transcript isoforms, leading to challenges in accurately quantifying the expression of different transcripts and interrogating alternative splicing patterns and differential isoform expression (0).

Biomarker discovery within the transcriptome can be extended beyond expression levels to include the detection of co-/post-transcriptional modifications such as 3’ end modification by addition of a polyadenine (poly(A)) tail facilitated by poly(A) polymerases (1). RNA Poly(A) tails play a role in post-transcriptional regulation, including mRNA stability and translational efficiency (2), where the length has been shown to be important in translation stimulation via poly(A) binding protein (PABP) (3). Furthermore, highly expressed transcripts have been shown to harbor shorter poly(A) tails (4). While poly(A) tail lengths have been investigated via head-to-tail ligation PCR (5) or alternative short-read sequencing techniques (e.g. PAL-seq (6) and TAIL-seq (7)), these homopolymers can extend to several hundred nucleotides (nt), which therefore poses limitations with short-read sequencing technologies (8). By design, short-read RNA-seq typically uses anchored oligo-dT priming for reverse transcription, which prohibits the capture of the full poly(A) tail length, failing to capture the full range of poly(A) tail lengths. Therefore, most biomarker discovery projects are unable explore co-/post-transcriptional modifications as potential biomarkers.

To overcome these challenges, an alternative strategy for RNA-seq has emerged, using direct or native RNA sequencing (RNA-seq) on an array of nanopores by Oxford Nanopore Technologies (ONT) (9-21). This advancement facilitates the direct analysis of RNA transcripts, minimizing potential errors and bias associated with cDNA synthesis and amplification, detection of polyadenylation length as well as the acquisition of long read data, which allows the identification of splice variants (0, 22), thus providing a more comprehensive view of the transcriptome. The additional information gained from this platform provides alternative methods of disease biomarker detection.

While the gene expression biases of Illumina cDNA sequencing have been widely studied, it remains unclear which biases are present in quantification of Nanopore direct RNA-seq, and whether Nanopore direct RNA-seq can be used in place of Illumina short-read sequencing in transcriptional biomarker discovery and validation studies (9-21). We therefore set out to compare blood mRNA data derived from patients with suspected viral or bacterial sepsis in previously published Illumina cDNA (3) with Nanopore direct RNA-seq data to understand the gene expression correlation between the two platforms. We also set out to investigate which additional information for biomarker studies could be obtained from Nanopore direct RNA-seq.

## RESULTS

### Comparison of gene and transcript expression quantification between direct RNA-sequencing and short-read Illumina cDNA-sequencing

To make comparisons between Illumina and Nanopore direct RNA-seq data, we sequenced RNA samples derived from whole blood of 12 patients with sepsis with Nanopore direct RNA-seq and compared the data to Illumina sequencing data (described in our previous work (3)). Nanopore sequencing yielded an average of 1,279,075 reads per sample (**Supplementary Table 1**). The aligned read lengths had a median of 971 nucleotides (**Supplementary Table 1**).

We evaluated the Pearson correlation in read counts per coding gene across different sequencing methods and all 12 samples (**Methods**). **Figure 1A** illustrates the correlations between Nanopore RNA-seq and Illumina cDNA-seq for all samples using widely-used RNA-seq quantification tools, including *NanoCount* (4), *IsoQuant* (5), *HTSeq* (6), and *Bambu* (7) for Nanopore RNA-seq, and *Kallisto* (8) and *HTSeq* (6) for Illumina RNA-seq (**Methods**). For the majority of individual samples, high correlations were observed between Nanopore and Illumina RNA-seq, with the highest correlations found between *NanoCount* and *Kallisto* (r=0.734-0.981, mean 0.927), followed by *IsoQuant* and *Kallisto* (r=0.695-0.983, mean 0.910), *HTSeq* and *Kallisto* (r=0.644-0.980, mean 0.885), *Bambu* and *Kallisto* (r=0.360-0.926, mean 0.760), and *HTSeq* and *HTSeq* (r=0.312-0.617, mean 0.500) (**Figure 1A**). Overall, we observed better consistency between *Kallisto* and other Nanopore RNA-seq tools compared to using *HTSeq* for both Nanopore and Illumina RNA-seq (**Supplementary** Figure 1), which suggested that *Kallisto* performed better than *HTSeq* for short-read sequencing performed on the Illumina platform. Additionally, we used Jensen–Shannon Divergence (JSD) to measure the similarity between the distributions of Nanopore RNA-seq and Illumina RNA-seq data for all samples using various RNA-seq quantification tools, where two identical distributions have JSD = 0 (the smaller, the better). *NanoCount* and *Kallisto* outperformed the alternatives, not only in terms of mean JSD values (mean 0.168) but also in their variances (**Figure 1B**). We further evaluated gene-to-gene correlations between Nanopore RNA-seq and Illumina RNA-seq and observed that the number of highly correlated genes increased as we excluded genes with low expression levels (**Supplementary** Figure 2A). A similar trend was noted for transcript-to-transcript correlations (**Supplementary** Figure 2B). Correlations between Nanopore RNA-seq (*Nanocount*) and Illumina RNA-seq (*Kallisto*) were lower (r = 0.435-0.885, mean = 0.736) compared to those observed at the gene level (**Supplementary** Figure 3).

**Figure 1.**
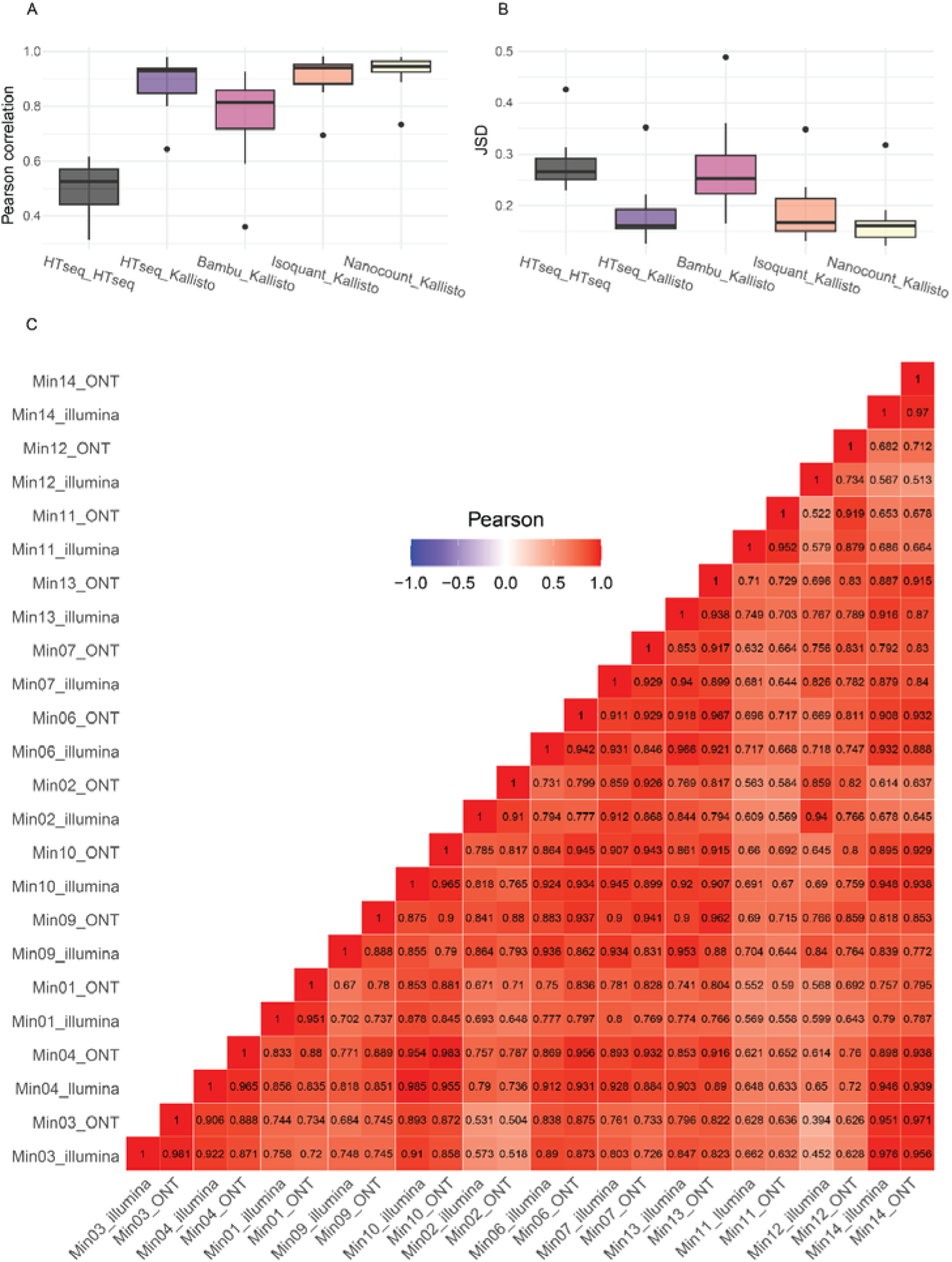
Gene-to-gene comparison with direct RNA sequencing and Illumina short sequencing using different pipelines. **A)** Pearson correlations between Nanopore RNA-seq and Illumina RNA-seq for all samples using different quantification tools, including NanoCount, IsoQuant, HTSeq, Bambu, and Kallisto. The order of the keys on the X-axis is ONT_Illumina, for example, HTSeq_Kallisto represents HTSeq for ONT correlated with Kallisto for Illunima. **B)** JSD (Jensen–Shannon Divergence) between Nanopore RNA-seq and Illumina cDNA-seq for all samples using different RNA-seq quantification tools, including NanoCount, IsoQuant, HTSeq, Bambu, and Kallisto. **C)** The heatmap of Pearson correlations on coding genes across all 12 samples using NanoCount for Nanopore and Kallisto for Illumina RNA-seq.

Interestingly, we noted that when analyzing Illumina data with *Kallisto* (8), the pipeline mitigated biases introduced by gene lengths by utilizing the Transcripts Per Million (TPM) metric (p > 0.37) (**Supplementary** Figure 4A). However, we observed a length bias in Nanopore data with *NanoCount*, even when using the same TPM metric (p < 0.00001) (**Supplementary** Figure 4B)., which may be reflective of more rapid sequencing of shorter reads. Furthermore, GC content impacted both *Kallisto* and *NanoCount* (p < 0.005) (**Supplementary** Figures 4C-D).

Collectively, our results highlight that gene expression estimates from Illumina and Nanopore platforms are highly correlated with certain combinations of pipelines, especially when using *NanoCount* for Nanopore direct RNA-seq and *Kallisto* for Illumina sequencing. Furthermore, length-dependent biases are more prevalent in Nanopore sequencing and GC content biases are present in both of sequencing platforms.

### Poly(A) tail lengths of mitochondrial vs non-mitochondrial transcripts in human blood mRNA

From the results above, it was apparent that since we obtained similar quantification outputs to Illumina cDNA-seq with Nanopore RNA-seq, the two platforms maybe considered equivalent terms of expression estimations, noting that Illumina cDNA-seq still remains more cost-effective. However, as mentioned previously, Nanopore direct RNA-seq provides additional advantages with its long-read capability, such as poly(A) tail length detection, although it remains unclear whether these features are important for biomarker discovery.

We therefore estimated the length of poly(A) tails at the 3’ end of transcripts using the built-in function of the ONT *Dorado* basecaller (**Methods**) (9). For mitochondrial transcripts, the overall distribution of poly(A) lengths was centred at ∼45 nt, and few poly(A) tails exceeded 70 nt in length (**Figure 2A**). These findings are consistent with previous studies on mitochondrial poly(A) RNA in human cell lines (0, 31). In contrast, nuclear transcripts exhibited a wider length distribution across the 12 samples, with a peak around ∼80 nt, and an average of 0.21% of poly(A) tails of transcripts across the samples were longer than 350 nt (**Figure 2B**). This highlighted the capability of long-read sequencing for transcriptome-wide poly(A) length estimations.

**Figure 2.**
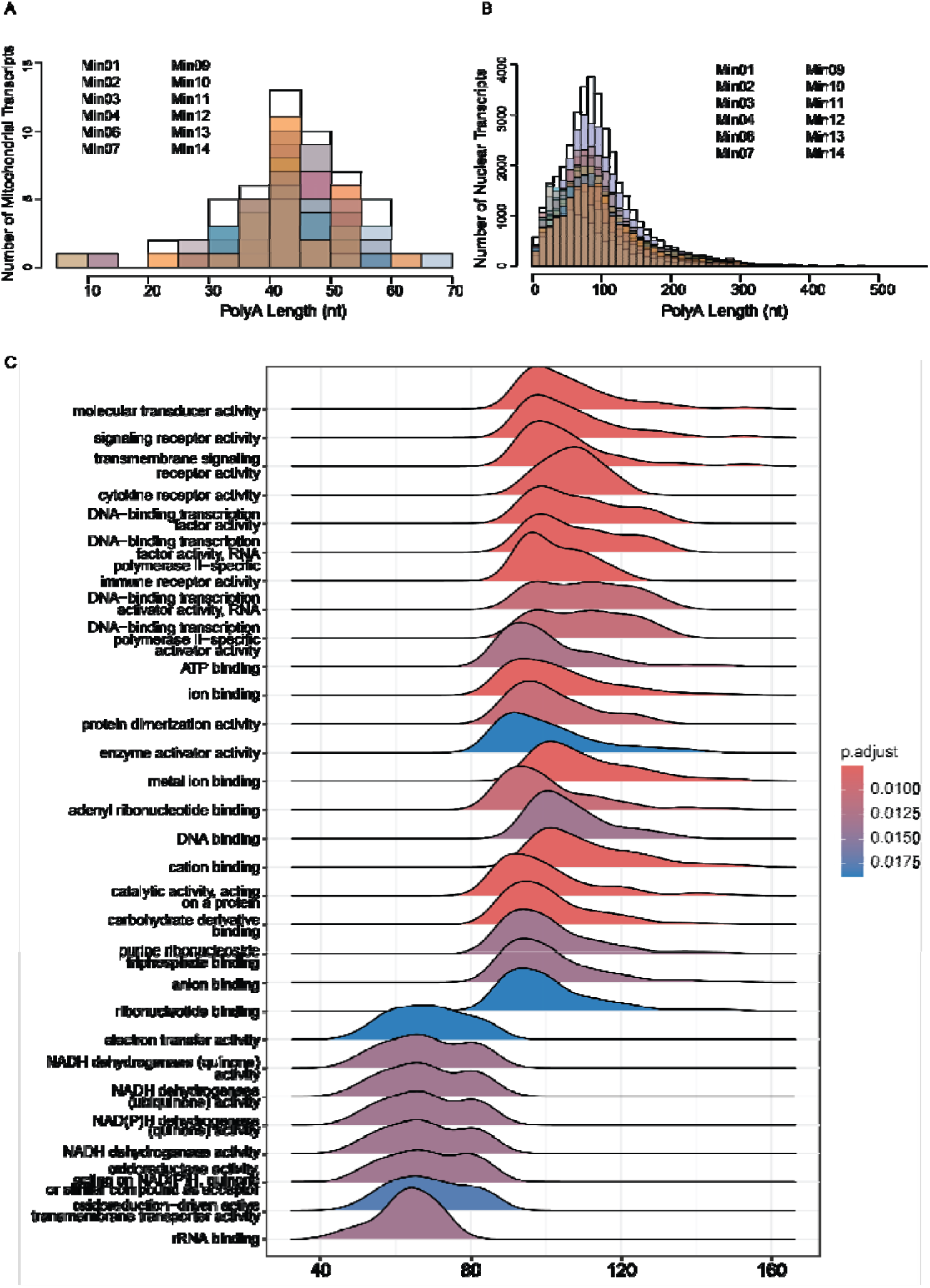
Poly(A) length distribution and Gene Set Enrichment Analysis (GSEA) using genes ranked by poly(A) tail lengths. **A)** Poly(A) length distribution in mitochondrial transcripts. **B)** Poly(A) length distribution in nuclear transcripts. **C)** Ridgeplot from the clusterProfiler bioconductor package with the X-axis indicate the poly(A) lengths. The distribution is the distribution of poly(A) length of those genes that enriched in the corresponding GO enrichment analysis (molecular function), and the colour indicates the significance, with adjusted P-value < 0.05 as significant (the full list of significant pathways can be viewed in **Supplementary Table 2**). The mitochondrial transcripts are excluded.

### GSEA of genes ranked by poly(A) tail lengths highlights molecular pathways enriched in genes with short and long poly(A) tails

Whether poly(A) tail lengths are randomly distributed or specific to functional units of cellular pathways is yet to be fully understood. Gene Set Enrichment Analysis (GSEA) identifies pathways where genes are enriched at the extremes of the ranked gene list, more than would be expected by chance alone. Traditionally, GSEA has found widespread application in the analysis of genes based on their differential expression rank or other scores (2-34). Here, we employed pre-ranked GSEA using the GSEA *R* packages on 1,520 coding genes, excluding mitochondrial transcripts (5, 36). In our study, genes were ranked according to their median poly(A) tail lengths, from longest to shortest. The median poly(A) tail lengths for the coding genes ranged from 26 nt to 147 nt, with a mean of 83 nt. We conducted GSEA to explore the Gene Ontology (GO) terms (**Figure 2C, Supplementary Figures 5-6, Supplementary Tables 2-4**) and Kyoto Encyclopedia of Genes and Genomes (KEGG) pathway databases (**Supplementary** Figure 7 **& Supplementary Table 5**) and identified pathways significantly associated with longer or shorter poly(A) tails.

The GO term analysis revealed that genes with shorter poly(A) tails exhibited significant enrichment in functions related to energy production and protein synthesis such as *oxidoreductase activity, NADH dehydrogenase activity, NAD(P)H dehydrogenase activity, oxidoreduction-driven active transmembrane transporter activity, electron transfer activity, large ribosomal subunit rRNA binding* and *structural constituents of ribosomes* (**Figure 2C**). The presence of shorter poly(A) tails in these pathways suggests that stability of mRNA derived from genes in these pathways may be reduced compared to genes belonging to other cellular pathways (7). In contrast, the recent evidence regarding abundant and efficiently translated mRNAs across eukaryotes having shorter poly(A) tail lengths may suggest that the genes involved in these pathways may have higher abundance and/or efficient translation (4).

Genes with longer poly(A) tails were significantly enriched in functional categories pivotal for more specialized and regulated cellular processes. These functions are predominantly related to signal transduction, including *signalling receptor activity, molecular transducer activity, and transmembrane signalling receptor activity*, as well as *ion binding, metal ion binding,* and *cation binding* (**Figure 2C**). Other enriched functions include *DNA-binding transcription factor activity* involved in transcriptional regulation and *immune receptor activity* related to an immune response. The longer poly(A) tails in these genes may enhance mRNA stability and translation efficiency, ensuring robust and sustained production of proteins involved in these complex and highly regulated pathways (8). In addition, we observed that the poly(A) distributions for each molecular functional pathway revealed a high degree of consistency across different samples. This consistency underscores the robustness of the poly(A) distribution patterns within each pathway, indicating that these distributions are maintained irrespective of sample variability (**Supplementary** Figure 8).

KEGG pathways belonging to infection, disease-related, ribosome and oxidative phosphorylation pathways comprised transcripts with shorter poly(A) lengths and immunity-related pathways showed longer poly(A) lengths overall (**Supplementary** Figure 7). This result suggests the potential stronger stability of immunity-related transcripts and high turnover of ribosomal and disease-related transcripts in patients experiencing an acute bacterial or viral infection Furthermore, we observed a bimodal distribution within one of the significant pathways - *insulin resistance* (**Supplementary** Figure 7). Upon investigating further, the first peak was enriched with a set of genes, including *SOCS3, TNFRSF1A, RPS6KA1, CD36, STAT3, PTEN, MLX*, and *PRKCB*. In contrast, the second peak notably included *PYGL* and *PPP1CB*, both exhibiting relatively longer poly(A) tails. *PYGL* and *PPP1CB* encode proteins that function as phosphatases, playing critical roles in metabolic regulation (9-42). Most genes in the first peak are actively involved in inflammatory processes and immune response, including *CD36* (3)*, SOCS3* (4)*, TNFRSF1A* (5)*, STAT3* (6)*, PTEN* (7), and *PRKCB* (8). These results highlight the importance of visualizing poly(A) tail lengths in RNA-seq data, as they may underlie diverse regulatory mechanisms and functional outcomes.

### Direct RNA sequencing uncovers hundreds of novel mRNA isoforms expressed in whole blood of patients with sepsis

Another advantageous feature of Nanopore sequencing is the ability to accurately determine novel transcript isoforms (9). Therefore, we explored novel isoform detection in our datasets. *IsoQuant* (5) has proven to be an effective tool for transcript discovery and quantification using long RNA reads, which showed correlation with Illumina cDNA sequencing comparable to *NanoCount* (**Figure 1**). We detected a total of 159,824 transcripts, of which 959 were considered novel isoforms by *IsoQuant*, with 594 non-artifact novel isoforms detected by *SQANTI3* after machine learning filtering (**Supplementary Table 6**). The majority of identified novel isoforms fell into the categories “Novel In Catalog”, “Incomplete-splice match” and “Novel Not in Catalog”, with an additional small fraction of query isoforms identified in other categories (**Figure 3A**). Overall, the set of novel transcript isoforms identified in Nanopore sequence data exhibited a wide range of inferred transcript lengths, from 340 to 8495 nt, with a mean length of ∼2199 nt across all categories (**Figure 3B**) and spanning all chromosomes (**Supplementary** Figure 9) with a peak on chromosome 1. Consistent with previous literature (0), the identified novel isoforms were often multi-exonic, with a mean exon count of 9.29 (**Figure 3C**). The gene with the most novel isoforms (9) was *IGLL5* (immunoglobulin lambda like polypeptide 5). These results highlight the potential to discover novel isoforms using Nanopore direct RNA sequencing on primary samples.

**Figure 3.**
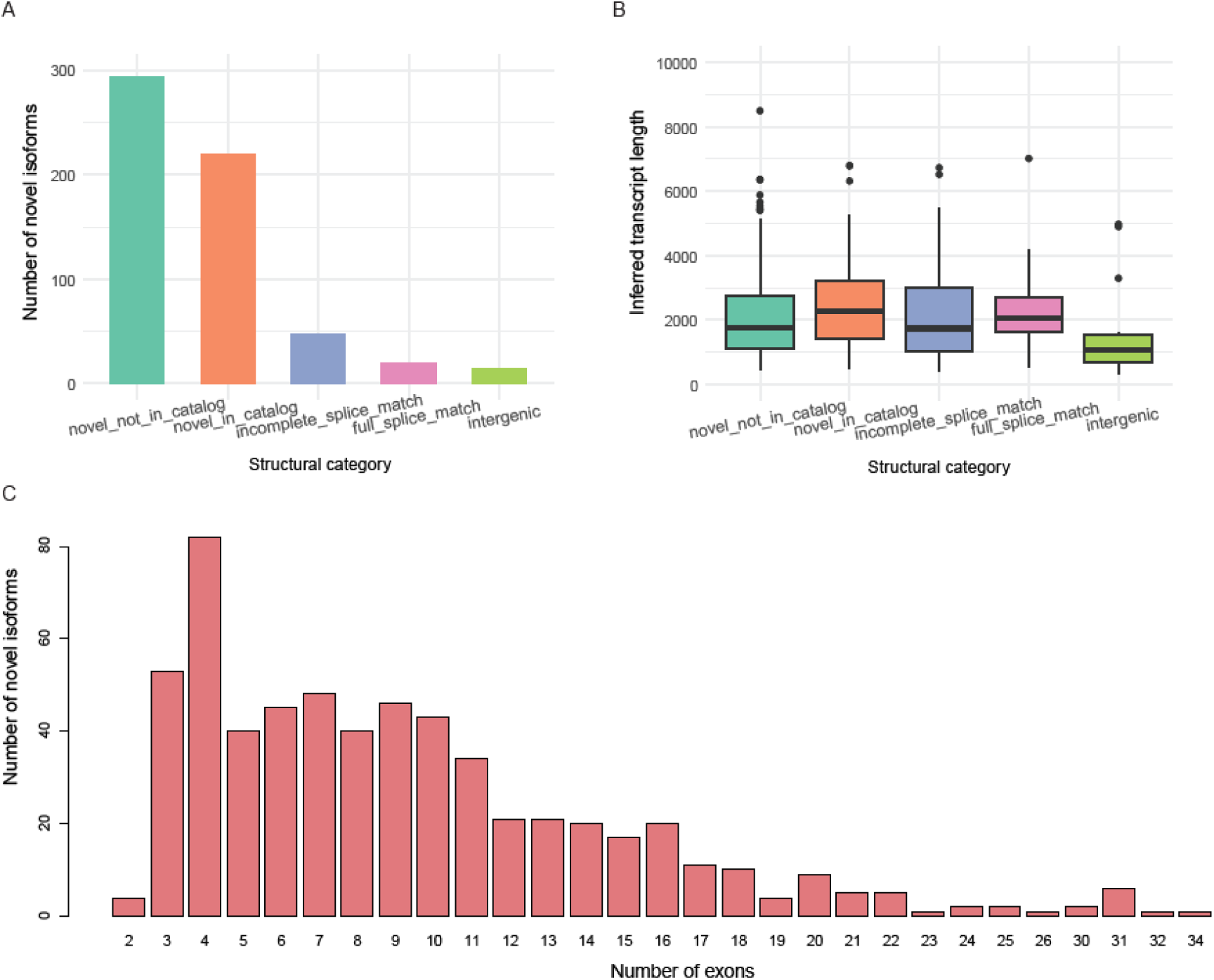
Characterization of novel isoforms identified by IsoQuant and SQANTI3. **A**) Structural category distribution for de novo identified isoforms. The structural category for an isoform indicates its relation to the closest annotated transcript. **B)** The length distribution of transcripts, stratified by the relation to the annotated transcripts (represented by the assigned structural category). The center line represents the median; hinges represent first and third quartiles; whiskers the most extreme values within 1.5 interquartile range from the box. **C)** The exon number distribution for identified isoforms.

### Investigating differential expression and polyadenylation between bacterial and viral infection

The samples we have studied here were a selected small subset of a larger study of 907 patients investigated via Illumina cDNA-seq for differences in host transcriptional response associated with confirmed bacterial or viral infection (3). The bacterial and viral pathogens detected in these samples is shown in **Supplementary Table 10**. It was of interest to see whether we could recapitulate the major differentially expressed genes identified in this larger comparison using Nanopore direct RNA-seq. To this end, we carried out a differential gene expression analysis between Nanopore direct RNA-seq data on 6 patients with definite bacterial infection and 6 patients with definite viral infection (**Methods**). A total of 9 significant differentially expressed genes (DEGs) were identified when applying thresholds of adjusted P-value < 0.05 and |logFC| ≥ 1. Of these, 8 DEGs were more highly expressed in patients with viral infection, while 1 was more highly expressed in patients with bacterial infection (**Figure 4A**). Notably, all these 9 DEGs were consistent with DEG results obtained from Illumina cDNA-seq, in our previous work (3). This consistency underscores the reliability and validity of our findings across different sequencing platforms.

**Figure 4.**
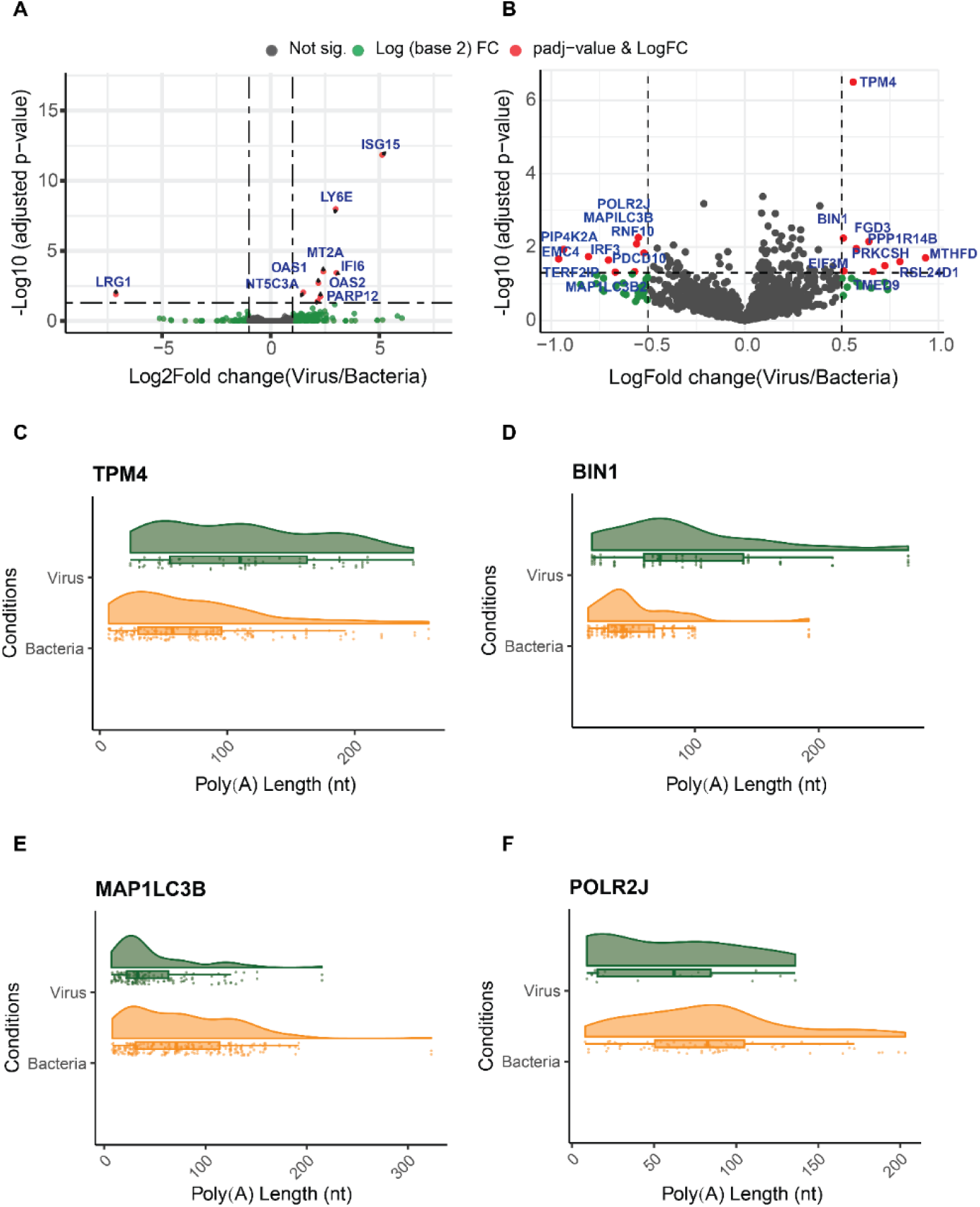
Differential expression and polyadenylation volcano plots. **A)** Volcano plots of viral vs bacterial differential expression from Nanopore direct RNA-seq datasets. Red dots indicate differentially expressed genes (DEGs) using adjusted P-value < 0.05 and |log2FC| ≥ 1 as cutoffs. **B)** Volcano plot of differential polyadenylation results from linear mixed-effects regression (lmer). Red dots indicate differentially polyadenylated genes (DPGs) using adjusted P-value < 0.05 and |logFC| ≥ 0.5 as cutoffs. **C-F)** Raincloud plots showing read-level polyadenylation estimates for top significantly differentially polyadenylated genes for **C)** TPM4 (adjusted P-value=3.20e-7), **D)** BIN1 (adjusted P-value=0.0057), **E)** MAP1LC3B (adjusted P-value=0.0082), and **F)** POLR2J (adjusted P-value=0.00551). Each point corresponds to a single read.

Following this, we focused on differential polyadenylation (DP) analysis using linear mixed-effects regression (*lmer*) (1) (**Methods**). Through the differential polyadenylation analysis of blood from 6 patients with viral infection and 6 patients with bacterial infection, using thresholds of adjusted P-value < 0.05 and |logFC| ≥ 0.5 (**Methods**), we identified 18 differentially polyadenylated genes (DPGs). Among these, 9 DPGs (*BIN1, EIF3M, FGD3, MTHFD2, PPP1R14B, PRKCSH, RSL24D1, TMED9*, and *TPM4*) exhibited increased polyadenylation, and 9 (*EMC4, IRF3, MAP1LC3B, MAP1LC3B2, PDCD10, PIP4K2A, POLR2J, RNF10*, and *TERF2IP*) exhibited decreased polyadenylation in the samples from patients with viral compared to bacterial infection (**Figure 4B, Supplementary Table 7**). Raincloud plots (2) were generated to visualize raw poly(A) tail lengths for all reads mapped to each of the DPGs under both conditions (viral and bacterial infections) (**Methods**). We present two genes that appeared to be most significantly differentially polyadenylated in **Figures 4C-4F**.

These observed differences showed more genes with DP than differential expression (DE), although with smaller effect sizes (**Figures 4A-B**). Overall, these results imply variations in the dynamic regulation of gene expression at the post-transcriptional level between viral and bacterial infections, and therefore, suggests the potential utility of polyadenylation as a disease biomarker.

### Investigating differential transcript usage between patients with confirmed bacterial and viral infection

Next, we explored differential transcript usage (DTU) - the variation in the proportion of different transcript isoforms per gene across different conditions - between blood samples from patients with viral and bacterial sepsis. Using *DRIMSeq* (3) and *StageR* (4), we observed significant DTU between viral and bacterial infection samples (**Methods**, **Supplementary Tables 8-9**). In total, four genes, *SOD2, RPS21, CD36*, and *RPL37*, showed significant DTU with adjusted P-value < 0.05 (**Supplementary Table 9**). For the gene *SOD2* (*ENSG00000112096.19*), transcript *ENST00000367055.8* (adjusted P-value = 0.029) showed reduced usage, whereas transcript *ENST00000538183.7* (adjusted P-value = 0.003) exhibited increased usage in samples from patients with viral compared to bacterial infection (**Figure 5A; Supplementary** Figure 10). Similar patterns of differential transcript usage were identified for the genes *RPS21* (*ENSG00000171858.18*), *CD36* (*ENSG00000135218.19*) and *RPL37* (*ENSG00000145592.14*). These genes were of interest as, *RPS21* and *RPL37* are both genes encoding ribosomal proteins (5), indicating the essential role of protein synthesis. *SOD2* is a critical regulator of antiviral signalling (6), while *CD36* is known to promote inflammatory responses and phagocytosis, processes involved in the host response to both viral and bacterial infections (3, 57, 58). For *RPS21*, transcript *ENST00000343986.9* (adjusted P-value = 0.010) showed increased usage, while *ENST00000450116.6* (adjusted P-value = 0.002) showed reduced usage (**Figure 5B**). For *CD36*, both transcripts, *ENST00000394788.7* and *ENST00000447544.7* (adjusted P-values = 0.000 for both), showed significant changes in usage (**Figure 5C**). Lastly, for *RPL37* (*ENSG00000145592.14*), only transcript *ENST00000504562.1* (adjusted P-value = 0.003) showed increased usage (**Figure 5D; Supplementary** Figure 10).

**Figure 5.**
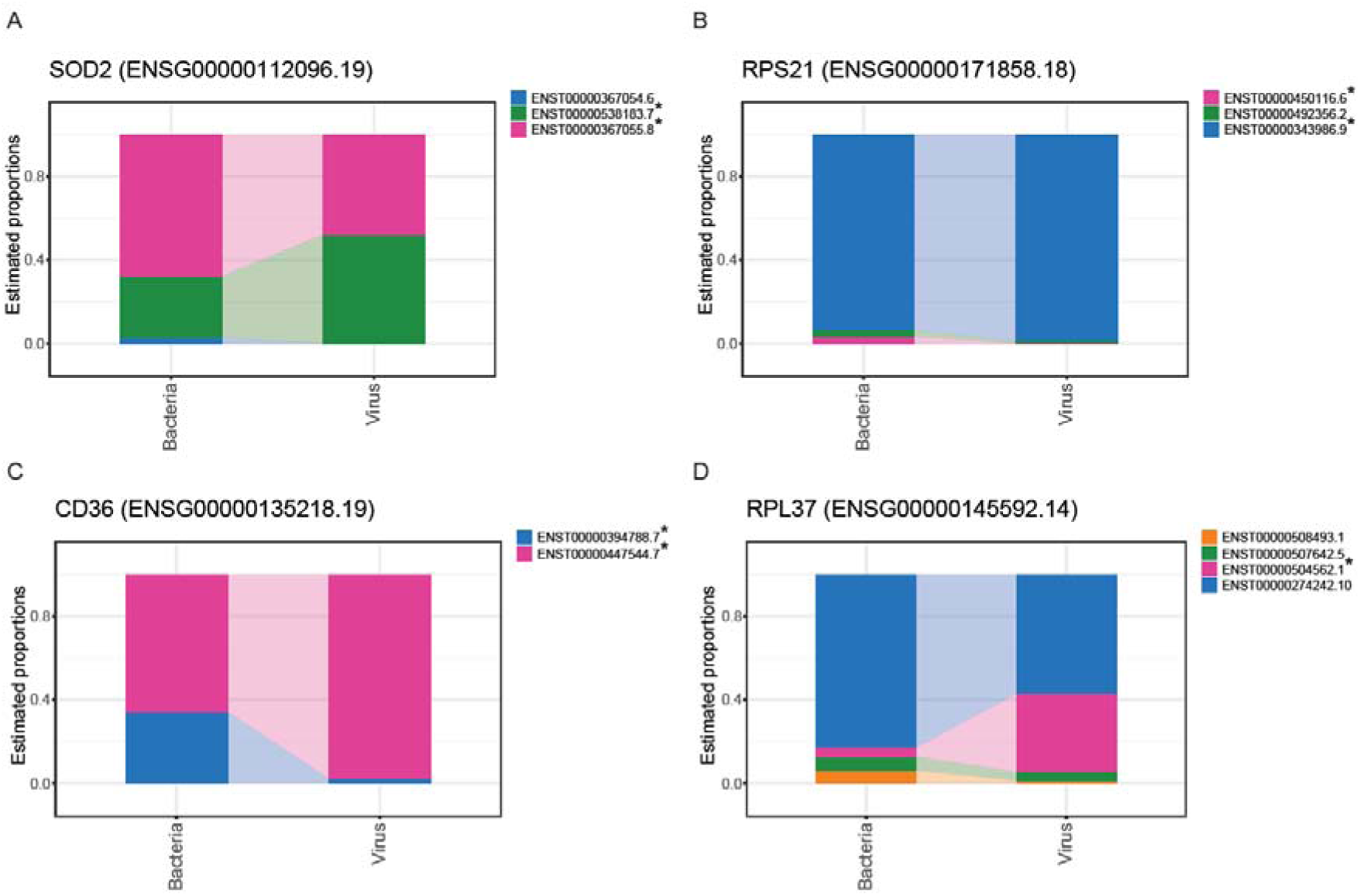
Differential transcript usage occurs between bacterial and viral samples. A-D. Differential estimated proportions of transcripts of genes for **A)** SOD2 (ENSG00000112096.19), B) RPS21(ENSG00000171858.18), **C)** CD36 (ENSG00000135218.19), and **D)** RPL37 (ENSG00000145592.14), with adjusted P-values < 0.05. Asterisks indicate transcripts which meet the adjusted P-value threshold of <0.05.

These findings highlight the utility of Nanopore RNA-seq in uncovering differences in the host response to bacterial and viral infection. By identifying both known and novel transcripts, this technology provides critical insights into pathogen-specific gene expression, which could be pivotal for understanding the molecular mechanisms underlying viral and bacterial infections.

## DISCUSSION

Nanopore direct RNA-seq has several advantages over other RNA sequencing approaches; **1)** the real-time nature of Nanopore sequencing expedites data acquisition and analysis; **2)** direct analysis of RNA molecules removes the need for cDNA sequencing, hence eliminates the bias introduced by cDNA preparation; **3)** it also enables continuous reads spanning many thousands of nucleotides, facilitating the identification of splice variants and novel transcript isoforms (0); and **4**) the unique 3’ priming method allows the full length detection of poly(A) tails on mRNA transcripts. While each of these features holds individual utility, their combination is unparalleled and promises to yield novel insights into RNA biology.

We first underscored a high level of agreement between Nanopore direct RNA-seq and Illumina cDNA-seq of mRNA levels within our blood mRNA samples, especially with the combination of *NanoCount* for Nanopore and *Kallisto* for Illumina sequencing (**Figure 1A**). Correlation analyses revealed concordance at the gene-to-gene levels (**Figures 1A-C**), indicative of the reliability and consistency of both technologies in capturing gene expression profiles. In short-read sequencing, the reads are often shorter than the transcripts they originate from, leading to multiple reads aligning consecutively to the gene locus in the reference genome. This can introduce a bias in measuring expression levels, as shorter transcripts may appear to be less expressed (1). Therefore, the high agreement levels at the gene-to-gene level were surprising. However, the transcript-level analysis showed that the correlations were lower (**Supplementary** Figure 3). This is more in line with our understanding that Illumina sequencing, with its shorter read lengths, is less effective at accurately capturing isoform level information, in which its biases and lack of correct transcript assignment would be exacerbated at the transcript level. However, surprisingly, when we explored gene length-dependent bias, analyzing Nanopore data with *NanoCount* showed evidence of gene-length bias when the TPM metric was used (p < 0.00001) (**Supplementary** Figure 4B). Long-read sequencing theoretically should reduce such biases, as a single long read can cover most of a transcript. This discrepancy may be due to Nanopore sequencing potentially overcounting shorter genes, as they pass through the Nanopore more quickly per read. Despite the significant correlation between gene-length and TPM, we note that the R^2^ value was low (R^2^ = 0.003). Therefore, improved RNA-seq quantification tools are needed to thoroughly address this issue and enhance the correlation between these two platforms.

Furthermore, we note that we observed better consistency between *Kallisto* and other Nanopore RNA-seq tools compared to *HTSeq* for Illumina data analysis (**Supplementary** Figure 1), and this is partly because *HTSeq* does not use a probabilistic model for ambiguous reads. Given the high rates of multi-mapping in RNA-seq data, the use of probabilistic models is crucial for achieving precise abundance estimates (2). Overall, further comparative assessments between Nanopore and Illumina RNA-seq expressions should be carried out to further examine these correlations with synthetic RNA with known concentrations, such as Sequins (3).

Variation in results from the same RNA-seq tool across different samples may arise from biological differences, such as varying gene expression levels or RNA degradation, as well as technical factors like sequencing depth or RNA quality. Sample complexity, including isoform diversity, can also contribute to variability in quantification. These factors can affect tool performance, leading to differences in transcript detection and abundance across samples (4, 65).

We utilised the unique capability of Nanopore RNA sequencing to explore polyadenylation in blood from patients with sepsis. The traditional understanding is that the average length of the poly(A) tail in mammalian mRNA is ∼100-250 nt, at the initial synthesis stage within the nucleus. However, upon length regulation of the poly(A) tail in the cytoplasm, the steady state length of the mRNA poly(A) tail has been identified to be shorter ∼50-100 nt (7, 66). In our study, our results agree with the idea that the average poly(A) tail length of non-mitochondrial transcripts in human blood mRNA is closer to ∼80 nt (**Figure 2B**). Furthermore, through subsequent GSEA based on poly(A) tail lengths, we identified specific pathways enriched with variations in polyadenylation. Interestingly, infection-, disease-related, ribosome- and oxidative phosphorylation-related pathways revealed to have shorter poly(A) lengths and immunity-related pathways such as *JAK-STAT signalling pathway* showed longer poly(A) lengths overall (**Supplementary** Figure 7). We note that particularly the group of KEGG pathways with shorter poly(A) tails such as *Parkinsons Disease*, *Huntington Disease*, *Oxidative Phosphorylation*, *Ribosome*, *Coronavirus disease - COVID-19*, are commonly enriched together in viral infections, such as SARS-CoV-2 infections (7-70). Considering our results were derived from patients with definite bacterial and viral infections, these findings shed light on the functional implications of altering poly(A) tail length in cellular functions, and the differential enrichment of poly(A) tail lengths across various biological pathways. Previously, transcripts with shorter poly(A) tails were shown to undergo faster rates of decay (6), which suggests the rapid regulation of these genes involved in the aforementioned pathways. Although it has been universally understood that longer poly(A) tails may lead to increased translation efficiency, a recent report suggests otherwise, where highly expressed and translated transcripts contain a shorter poly(A) tail (4). As it stands, the relationship between poly(A) length, expression and translation are still unclear and will need further investigations. Furthermore, the number of DPGs outweighed the number of DEGs between viral and bacterial samples (**Figures 4A-B**). Through this result, we highlight the potential of polyadenylation as a plausible method of biomarker discovery for disease.

Our study also revealed numerous novel isoforms through Nanopore direct RNA-seq (**Supplementary Table 6**), highlighting the utility of long-read sequencing in discovering novel transcripts. The identified isoforms exhibited a diverse array of characteristics and were associated with various biological processes, underscoring the complexity and heterogeneity inherent in the transcriptome. Notably, several novel isoforms were linked to genes implicated in immune-related pathways and diseases, hinting at their potential roles in pathophysiology. Nine novel isoforms were identified for *IGLL5* (immunoglobulin lambda like polypeptide 5), which has been reported to be correlated with tumor - infiltrating immune cells (1) and implicated in Multiple myeloma (2) and mature B-cell lymphoma (3). Continued efforts to understand the diversity of the transcriptome is crucial in identifying causes and treatment options for disease, and novel isoform discovery is one promising and important method of improving our understanding. As only long-read sequencing can capture the full lengths of transcripts, and therefore identify splicing patterns accurately within isoforms, we expect that Nanopore or Pacific Bioscience (PacBio) will be utilized as gold standards for transcript isoform discovery.

Lastly, we identified significant differential transcript usage (DTU) for several genes between viral and bacterial samples using both known and novel transcripts from Nanopore RNA-seq (**Figure 5**). While differential gene expression is widely used in RNA-seq studies, DTU explores the transcriptome at the transcript/isoform-level and is less frequently studied. This approach provides crucial insights into pathogen-specific gene expression, which are essential for understanding the molecular mechanisms underlying viral and bacterial infections. For instance, our data analysis revealed only 9 significant DEGs (**Figure 4A**), but we were able to further interrogate the transcriptomic changes by visualizing the DTU at the gene level and isoform level (**Figure 5**), which also highlights the potential of DTU being used for biomarker detection for disease states.

There are, however, some shortcomings associated with Nanopore direct RNA-seq (8, 74, 75) in comparison with Illumina cDNA-seq. The throughput of Nanopore direct RNA-seq remains lower than that of other high-throughput sequencing platforms, such as Illumina cDNA-seq, potentially limiting its use in large-scale studies (4, 75). Also, most available and established pipelines have been designed and tested for Illumina cDNA-seq, whereas most Nanopore RNA pipelines are newly developed by the user community and are less maintained and kept up to date in comparison. Furthermore, input requirements for Nanopore sequencing is much higher than that of Illumina cDNA-seq, especially with direct RNA-seq protocols. Although recent developments in direct RNA-seq have allowed for lower input requirements, the lack of a PCR step in the protocol means that for precious or low-yield samples, e.g. clinical samples, ONT direct RNA-seq may not be feasible.

This current study has various limitations. We have explored a small number of samples (6 in each condition – bacterial vs viral infection), and the results of our statistical analyses will be enhanced by incorporating an increased number of samples. Furthermore, a major advantage of Nanopore direct RNA-seq is the ability to direct post-transcriptional modifications such as nucleotide modifications (1), which we did not explore within this study. RNA modification analysis tools are rapidly evolving and being developed at unprecedented rates, with many variations in outcomes and there is currently no gold standard method for understanding RNA modifications with direct RNA-seq. Currently, the newest versions of the ONT basecaller *Dorado* can detect RNA modifications during the basecalling for the updated direct RNA-seq kit (SQK-RNA004, whereas this current study utilizes the older kit SQK-RNA002), which has exponentially increased the ease of analyzing the modifications. We expect that with further improvements to the *Dorado* algorithm, accurate and rapid detection of modifications will be possible, which would lead to the potential use of this technique for biomarker detection, as we have discussed with polyadenylation and DTU.

## CONCLUSIONS

Our comparison of the two sequencing technologies - ONT direct RNA-seq and Illumina cDNA-seq - demonstrates that, with the application of a well-optimized analysis pipeline, there is a strong correlation between gene expression estimates derived from both Illumina and Nanopore platforms. However, ONT direct RNA sequencing offers unique advantages not provided by Illumina cDNA sequencing. Notably, Nanopore RNA-seq reveals critical aspects of RNA regulation, such as variations in poly(A) tail length and the discovery of novel isoforms, which are not easily detectable through Illumina cDNA sequencing. Additionally, our analysis identifies significant variations in poly(A) tail length that are closely related to molecular functions, offering a deeper understanding of gene expression and its regulatory mechanisms. Our results suggest that integrating Nanopore direct RNA sequencing into research workflows could significantly enhance insights into RNA regulation and gene expression, providing valuable contributions to understanding disease mechanisms.

## METHODS

### Study design and participants

The samples in this study were selected from RNA collected for a larger study of 907 children evaluated for sepsis (3). Bacterial infections were confirmed by cultures of sterile sites by standard pathology services which must be compatible with the clinical presentation. Confirmed viral infection were based on routine diagnostics (influenza A and B, respiratory syncytial Virus (RSV), parainfluenza 1-3, human metapneumovirus (hMPV), adenovirus, enterovirus) and add-on viral diagnostics of specimens as clinically indicated (such as Enterovirus-PCR in infants with suspected sepsis or central nervous system infection).

### Sample Collection and Processing

Blood samples were collected from children with suspected sepsis. 2.5mL of blood was collected in PAXgene Blood RNA tubes (PreAnalytix) and total RNA was extracted using PAXgene Blood miRNA Kit (PreAnalytix).

### RNA QC and Quantification

RNA samples were quantified using the Qubit™ RNA broad range Assay Kit (Invitrogen) and QC was performed using the Agilent RNA assay (#5067-5576) on the TapeStation 4200 (Agilent # G2991AA) as per the manufacturer’s protocol.

### GLOBINclear™-Globin mRNA Depletion

1-4 μg of total RNA in a maximum volume of 14 μL was used to remove globin mRNA using the GLOBINclear™-Human Kit, for globin mRNA depletion kit (Invitrogen #AM1980), as per the manufacturer’s protocol. On completion of the mRNA depletion protocol, each RNA was quantified, and QC was performed using the Qubit™ RNA Broad Range Assay Kit (Invitrogen) and the Agilent RNA assay (#5067-5576) on the TapeStation 4200 (Agilent #G2991AA) as per the manufacturer’s protocol.

### ONT Library Preparation and Sequencing

Libraries were prepared following the Direct RNA Sequencing protocol (ONT, #SQK-RNA002) as per the manufacturer’s protocol. The only alteration to the protocol was the amount of input RNA to maximise the concentration of input RNA and accommodate samples with lower concentration (0 – 700 ng of globin-depleted total RNA). For samples with RNA concentration lower than 50 μg/μL, a maximum input volume of 9 μL was used to prepare the libraries. On completion of the library prep, the reversed-transcribed and adapted RNA was sequenced on a MinION Mk1B (Oxford Nanopore) using a R9.4.1 flow cell using *MinKNOW* v22.12.7 with the default settings when the flow cells was used once, and v20.06.18 with the default settings for a total of 24 hours if the flow cell was washed and re-used. On completion of the first round of sequencing, a flow cell wash was performed using a Flow Cell Wash Kit (ONT, #EXP-WSH004) as per the manufacturer’s protocol. Once the flow cell has been washed and pore QC checked, a second library was loaded and sequenced according to the same settings that was mentioned previously.

### Illumina cDNA-Sequencing

Data from Illumina cDNA-seq was derived from our previous work (3). Briefly, libraries were prepared from total RNA using the TruSeq Stranded Total RNA (RiboLJZero GOLD) Library Preparation kit (Illumina). Strand-specific libraries were sequenced using the Illumina NextSeq 75 cycle (1x75bp) High Output Run.

### Basecalling and Alignment

For the 12 Nanopore sequencing data, *Dorado* v5.3 was used for basecalling while the model was set as rna002_70bps_hac@v3 and “--estimate-poly-a” parameter was applied. Fast5 files were converted to Pod5 format before inputting to dorado software using “pod5 convert fast5” as per recommended in *Dorado* user manual. Only passed reads were kept for anlaysis. Bam format was set as the *Dorado* output format to keep more information including poly(A) tail length. By using “samtools bam2fq”, bam output files were converted to fastq format for mapping purpose. For 12 Illumina sequences, fastq files were demultiplexed on the sequencing machine. Gencode GRCh38 v35 transcriptome genome and GRCh38 full genome human reference was provided as the reference transcriptome genome and full genome reference when mapping the fastq files. *Minimap2 v2.24* was used for mapping, with command “minimap2 -t 20 -ax splice -uf -k14 -L ref.fa sample.fastq” for ONT sequences and default minimap2 short read parameter for Illumina sequences. *Samtools v1.16.1* was used to sort and index the bam file created from mapping process. Mapping statistic results were calculated using “samtools flagstats” function.

### Pearson Correlation Analysis of Nanopore and Illumina Sequencing Data

To evaluate the correlation between Nanopore direct RNA sequencing data and Illumina cDNA sequencing data, we conducted Pearson correlation analysis using *R*. Nanopore direct RNA-seq data underwent processing with various software packages, including *NanoCount* (4), *IsoQuant* (5), *HTSeq* (6), and *Bambu* (7) for Nanopore RNA-seq, a while Illumina cDNA sequencing data were processed with *Kallisto* (8) and *HTSeq* (6) for Illumina RNA-seq (**Methods**).

*Python3* and *R* scripts were developed to standardize transcript IDs and gene names across different software. Detailed instructions for using each software and their respective scripts can be found in their software documentation. Transcripts isoforms were grouped into genes using established gene annotation databases - Ensembl and GENCODE (6, 77).

For each combination of sequencing platform (Nanopore or Illumina) and processing software, raw count data or transcript-level abundance estimates were obtained. Pearson correlation coefficients were then computed between corresponding gene-to-gene expression values across samples for the mapped data. Transcript-level analyses were carried out without mapping to genes, but via calculating Pearson correlations directly. All correlation analyses were conducted in *R* v4.3.1, utilizing built-in functions for calculating Pearson correlation coefficients.

### Poly(A) Tail Length Analysis

When using the *Dorado* basecaller (9) with the parameter “--estimate-poly-a”, the output bam file will contain an extra tag to record the poly(A) tail length in each read. A summary on read length and poly(A) tail length was created with the command “samtools view basecalling.bam | awk ’/pt:i/{print $1,length($10),$NF}’ | sed ’s/pt:i://g’ “.

### Gene Set Enrichment Analysis (GSEA)

Gene Set Enrichment Analysis (GSEA) was conducted to identify significantly enriched pathways and biological processes associated with the experimental conditions. We utilized pre-ranked GSEA with the GSEA *R* packages, focusing on coding genes, excluding mitochondrial transcripts. Typically, GSEA is employed to analyze genes based on their differential expression ranks or other relevant scores. In our study, genes were ranked according to their poly(A) tail lengths, from longest to shortest.

For the analysis, we utilized the *clusterProfiler* package from Bioconductor. Specifically, the bridgeport function within *clusterProfiler* was used to perform the GSEA targeting the Kyoto Encyclopedia of Genes and Genomes (KEGG) pathways and Gene Ontology (GO) terms databases. For KEGG pathway enrichment analysis, the enrichKEGG function was utilized to identify significantly enriched pathways. Similarly, for GO term enrichment analysis, the enrichGO function was used to determine significantly enriched biological processes (BP), molecular functions (MF), and cellular components (CC). Enrichment scores and significance levels were computed using permutation testing, with a False Discovery Rate (FDR) threshold set at 0.05 to determine statistically significant enrichment. All analyses were conducted in *R* v4.3.1 with *clusterProfiler* v4.12.0 (6), ensuring reproducibility and robustness of the results.

### Differential expression Analysis

*DESeq2 v1.42.0* was used to identify differentially expressed genes from direct RNA-seq data. A minimum expression threshold of 10 reads per gene across all samples was applied. Comparisons between viral and bacterial infection samples were conducted using the standard pipeline. Genes with an adjusted P-value < 0.05 and |log2FC| ≥ 1 were considered significantly differentially expressed. Volcano plots were generated using the *EnhancedVolcano v1.20.0* package in R.

### Differential polyadenylation analysis

The differential polyadenylation analysis aimed to identify variations in poly(A) tail lengths across different experimental conditions, specifically comparing viral and bacterial infection samples. This analysis sought to elucidate how changes in polyadenylation patterns might correlate with gene expression and functional outcomes.

Poly(A) tail length measurements were obtained from Nanopore RNA-seq data, providing high-resolution insights into polyadenylation dynamics. The raw poly(A) lengths were log-transformed due to their right-skewed distribution. Subsequently, the package *lmerTest* v3.1.3 (1) was employed to perform a linear mixed-effects regression (*lmer*), where the log-transformed poly(A) length for all reads mapped to one gene served as the response variable, the infection type (viral or bacterial) as the fixed effect, and the sample batch as the random effect. Per-gene P-values were generated and adjusted using the Benjamini-Hochberg (BH) method with the ‘p.adjust’ function in R. Genes exhibiting differential polyadenylation were identified using cutoffs of an adjusted P-value < 0.05 and |logFC| ≥ 0.5.

Raincloud plots were generated for raw poly(A) tail lengths of all reads that mapped to each DPG under both conditions (viral and bacterial infections) using *ggplot2* v3.5.1, replicating the raincloud plots generated by the *raincloudplots* v0.2.0 package in *R* (2).

### Novel isoform identification

Due to low sequence coverage per sample, we aggregated the direct Nanopore RNA-seq data from all samples to detect novel isoforms. In addition to the novel transcripts discovered by *IsoQuant*, we applied *SQANTI3* (8) to the *IsoQuant* output GTF file containing the entire reference annotation plus all discovered novel transcripts. *SQANTI3* uses a random forest classifier to filter out artifacts by learning high and low-quality attributes from a True Positive (TP) and True Negative (TN) transcript set, building a model to distinguish artifacts and isoforms based on TN and TP properties (**Methods**).

*IsoQuant* was employed to discover novel transcripts using all 12 ONT datasets as input. Additionally, *IsoQuant* performed gene and transcript counting on both known and novel transcripts. Using the updated annotation gtf extended by *IsoQuant*, we ran the novel isoform discovery tool *SQANTI3* using the ‘qc’ function. Then, we used *SQANTI3* ‘filter’ to apply a machine learning classification method, with the default cut off threshold, to remove artifactual isoforms in the discovery output. Furthermore, we extended the annotation with new novel isoforms discovered from *IsoQuant* and *SQANTI3* and ran the *Featurecounts* tool to confirm the existence of the novel isoforms. *SQANTI3* classifies each isoform by finding the best matching reference transcript, in the following order [https://github.com/ConesaLab/SQANTI3/wiki/SQANTI3-isoform-classification:-categories-and-subcategories]:

1. FSM (Full Splice Match): meaning the reference and query isoform have the same number of exons and each internal junction agree. The exact 5’ start and 3’ end can differ by any amount.
2. ISM (Incomplete Splice Match): the query isoform has fewer 5’ exons than the reference, but each internal junction agree. The exact 5’ start and 3’ end can differ by any amount.
3. NIC (Novel In Catalog): the query isoform does not have a FSM or ISM match, but is using a combination of known donor/acceptor sites.
4. NNC (Novel Not in Catalog): the query isoform does not have a FSM or ISM match, and has at least one donor or acceptor site that is not annotated.
5. Antisense: the query isoform does not have overlap a same-strand reference gene but is anti-sense to an annotated gene.
6. Genic Intron: the query isoform is completely contained within an annotated intron.
7. Genic Genomic: the query isoform overlaps with introns and exons.
8. Intergenic: the query isoform is in the intergenic region.

### Differential Transcript Usage Analysis

We integrated the identified novel transcripts into the input annotation file and subsequently re-ran *IsoQuant*. Counts (TPM) derived from *IsoQuant* utilizing transcriptome-mapped BAM files were used to quantify the differential transcript usage between bacterial and viral samples. For differential transcript usage analysis, the quantified counts were input into *DRIMSeq* v1.14.0 (3), a tool designed to detect differences in transcript isoform usage. Prior to analysis, the counts underwent filtering based on specific conditions to ensure robustness and reliability. Specifically, parameters including *min_samps_gene_expr, min_samps_feature_expr, min_gene_expr, and min_feature_exp*r were set to 12, 4, 10, and 10, respectively. The output was used for stage-wise analysis using *StageR v1.26.0* (4), where the final list of significant genes and transcripts was filtered by adjust-P value < 0.05.

## Data Availability

The original datasets used and/or analysed during the current study will be uploaded to public repositories upon publication of this manuscript. Other data generated during this study are included in this published article and its supplementary information files.

## LIST OF ABBREVIATIONS

BH: Benjamini-Hochberg
BP: Biological processes
CC: Cellular components
cDNA-seq: cDNA-sequencing
DE: Differential expression
DEG: Differentially expressed gene
DP: Differential polyadenylation
DPG: Differentially polyadenylated gene
DTU: Differential transcript usage
FDR: False discovery rate
G-C: Guanine-cytosine
GO: Gene Ontology
GSEA: Gene Set Enrichment Analysis
JSD: Jensen–Shannon Divergence
KEGG: Kyoto Encyclopedia of Genes and Genomes
LogFC: Log fold change
MF: Molecular functions
nt: Nucleotide
ONT: Oxford Nanopore Technologies
Poly(A): Polyadenine
PCR: Polymerase chain reaction
RNA-seq: RNA-sequencing
TP: True positive
TPM: Transcripts per million
TN: True negative

## DECLARATIONS

### Ethics approval and consent to participate

The institutional Human Research Ethics Committee approved the study on June 9, 2017 (HREC/17/QRCH/85). Written informed consent or delayed consent was obtained for all participants from their parents/carers.

### Consent for publication

Not applicable.

### Availability of data and materials

The processed sequence data such as count matrix are available from https://github.com/abcdtree/dRNA-ONT-blood. The unprocessed sequence data will be made available via controlled access upon publication of this manuscript.

## Authors’ contributions

JH, DG, and LJMC developed the methodology and designed parts of the study. DG, ST, SN, HL, AB, SJM, JCK, and LJS conducted wet-lab experiments and generated the data for analysis. JH and JZ carried out the data analysis. JH, DG and LJMC contributed to the first draft of the manuscript. JH, DG, JJYC, SLT, SN, HL, AB, KSG, LJS and LJMC were involved in reviewing and editing the manuscript. All authors contributed to the article and approved the submitted version.

Rapid Paediatric Infection Diagnosis in Sepsis (RAPIDS) study group: Luregn J. Schlapbach^4,6^, Sainath Raman^4,9^, Natalie Sharp^4^, Natalie Phillips^4,10^, Adam Irwin^11,12^, Shane George^4,13,14^, Keith Grimwood^14,15^, Peter J. Snelling^13,14^, Arjun Chavan^16^, Allison Hempenstall^17^, Kristen S. Gibbons^4^, Renate Le Marsney^4^, Antje Blumenthal^4^, Devika Ganesamoorthy^2^, Lachlan Coin^1,3,7,9^

9 Paediatric Intensive Care Unit, Queensland Children’s Hospital, Brisbane, Australia.

10 Emergency Department, Queensland Children’s Hospital, Children’s Health Queensland, Brisbane, QLD 4101, Australia.

11 Faculty of Medicine, UQ Centre for Clinical Research, University of Queensland, Brisbane, QLD 4029, Australia.

12 Infection Management and Prevention Services, Queensland Children’s Hospital, Children’s Health Queensland, Brisbane, QLD 4101, Australia.

13 Department of Emergency Medicine, Gold Coast University Hospital, Southport, QLD 4215, Australia.

14 School of Medicine and Dentistry, Griffith University, Southport, QLD 4222, Australia.

15 Department of Infectious Disease and Paediatrics, Gold Coast Health, Southport, QLD 4215, Australia.

16 Paediatric Intensive Care Unit, Townsville University Hospital, Townsville, QLD 4814, Australia.

17 Thursday Island Base Hospital, Thursday Island, QLD 4875, Australia.

## Acknowledgements

We are grateful to all the parents and children participating this study, and the clinical and research teams who contributed to study setup, recruitment of patients, data collection and entry and monitoring. We thank the scientific teams for performing the RNA preparation and sequencing. We acknowledge all investigators of the Rapid Pediatric Infection Diagnosis in Sepsis (RAPIDS) Study Group, listed in the table below.

---

**RAPIDS Study Group** investigators, listed by study site:

| *Site* | **First Name** | **Family Name** | **Title** |
| --- | --- | --- | --- |
| *Queensland Children's Hospital* | Luregn J | Schlapbach | MD, PhD |
|  | Sainath | Raman | MBBS, PhD |
|  | Nathalie | Sharp | BSc |
|  | Natalie | Phillips | MBBS, MPhil |
|  | Adam | Irwin | MBCChB, PhD |
|  | Ross | Balch | PhD |
|  | Amanda | Harley | PhD |
|  | Kerry | Johnson | BSc |
|  | Zoe | Server | BSc |
| *Gold Coast University Hospital* | Shane | George | MBBS, MPH |
|  | Keith | Grimwood | MBCChB, MD |
|  | Peter J | Snelling | MBBS, MPHTM,<br>PhD |
| *Townsville University Hospital* | Arjun | Chavan | MBBS, MD |
|  | Eleanor | Kitcatt | MBBS |
|  | Luke | Lawton | MBBS |
| *Thursday Island Hospital* | Allison | Hempenstall | MD, MPH |
|  | Pelista | Pilot | - |
| *The University of Queensland* | Kristen S | Gibbons | PhD |
|  | Renate | Le Marsney | MPH |
|  | Antje | Blumenthal | Dr. rer. nat |
|  | Devika | Ganesamoorthy | PhD |
|  | Carolyn | Pardo | BSc |
|  | Jessica | Kling | MD, PhD |
|  | Stephen | McPherson | PhD |
|  | Anna D | MacDonald | PhD |
|  | Seweryn | Bialasiewicz | PhD |
|  | Trang | Pham | MEng |
| *The University of Melbourne* | Lachlan | Coin | PhD |

## SUPPLEMENTARY INFORMATION

Additional File 1: Figures. Supporting supplementary figures 1-12.

Additional File 2: Tables. Supporting supplementary tables 1-10.

