## Supplementary Figures for "Utilising Nanopore direct RNA sequencing of blood from patients with sepsis for discovery of co- and post-transcriptional disease biomarkers"

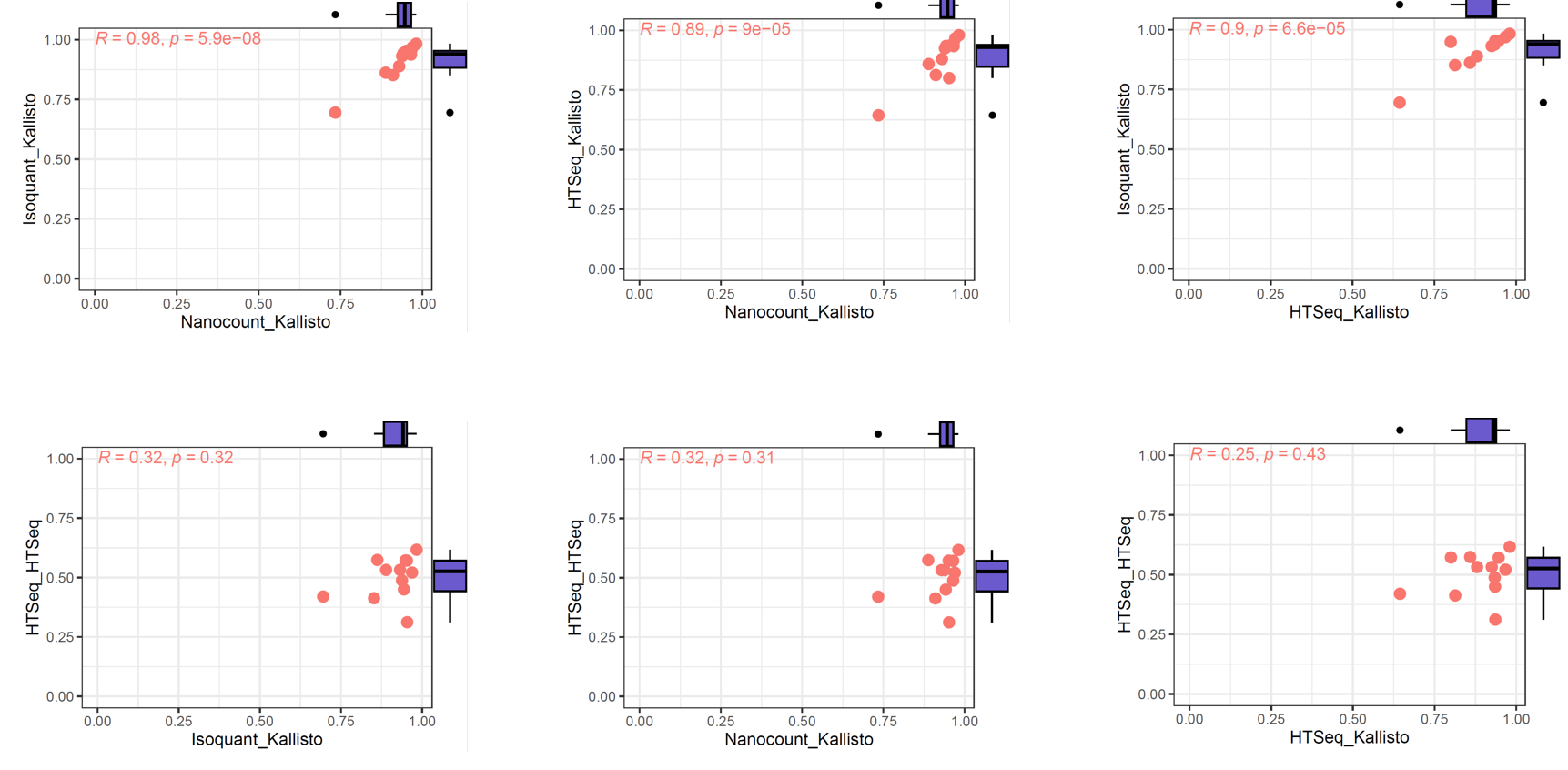

**Supplementary Figure 1. The correlation plots for each pair of software comparisons between Nanopore direct RNA-seq and Illumina cDNA sequencing.**

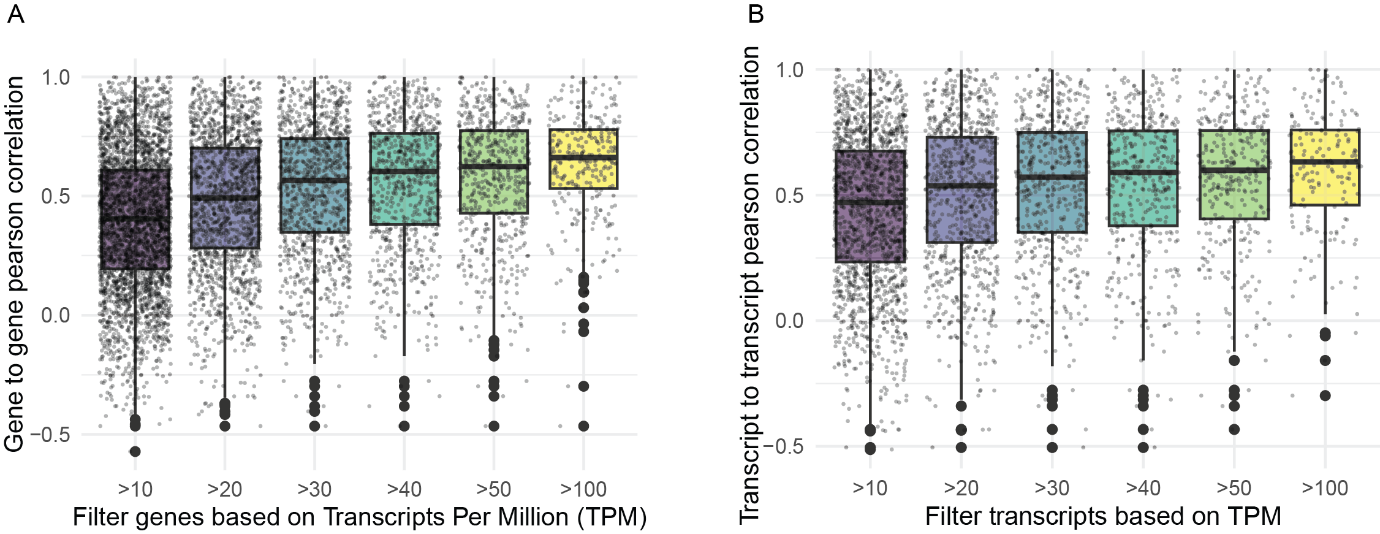

**Supplementary Figure 2. Comparison with Direct RNA-seq and Illumina cDNA-seq of matching 12 samples using different pipelines.** *NanoCount* for Nanopore and *Kallisto* for Illumina sequencing data were implemented, where Pearson correlations with **A)** gene-to-gene and **B)** transcript-to-transcript are shown. The X-axis indicates filter thresholds of the genes based on the level of TPM, and the Y-axis indicates the Pearson correlations.

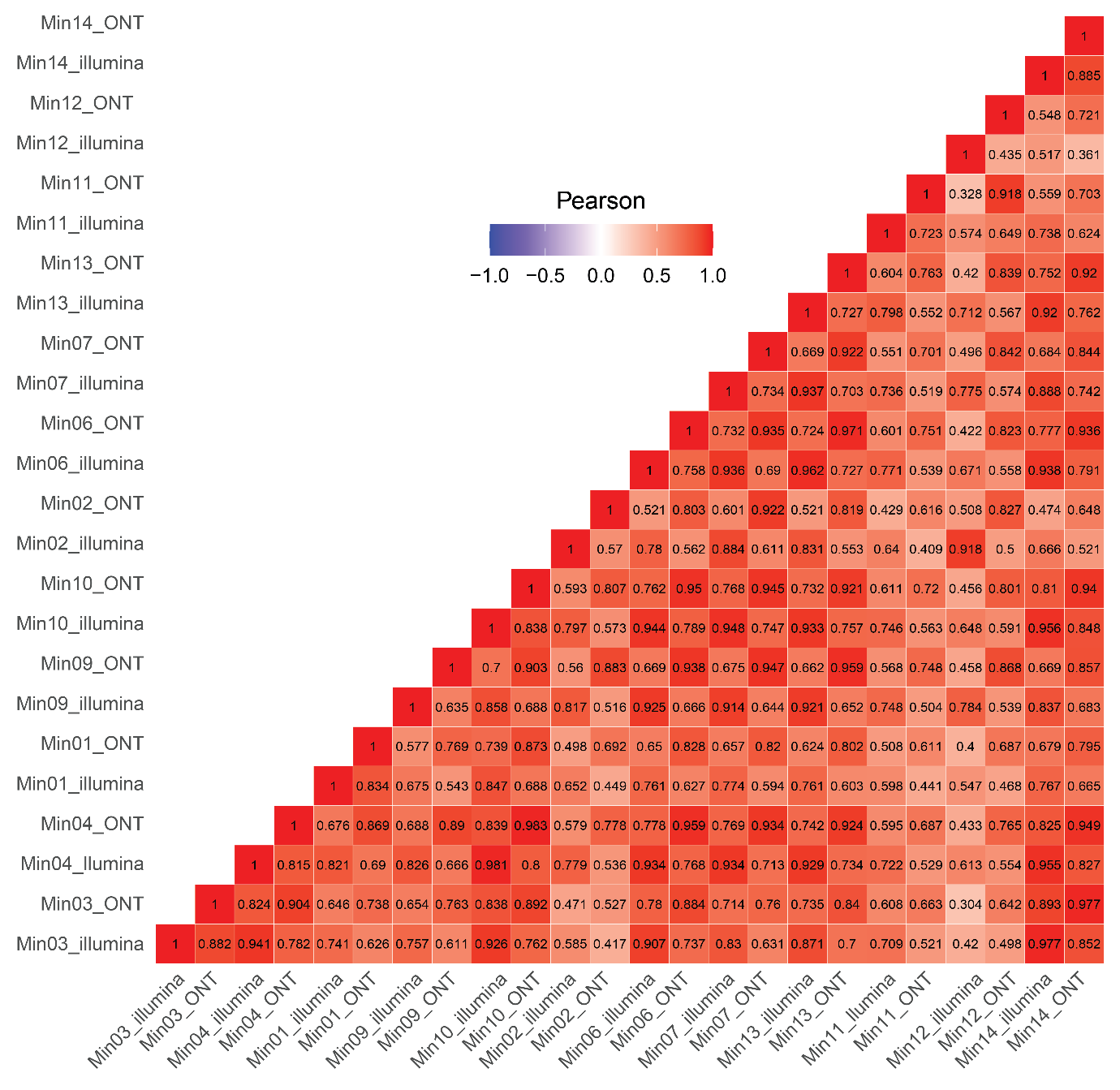

**Supplementary Figure 3. The heatmap of Pearson correlations on coding genes at the transcript level across all 12 samples using *NanoCount* for Nanopore and *Kallisto* for Illumina cDNA-seq.**

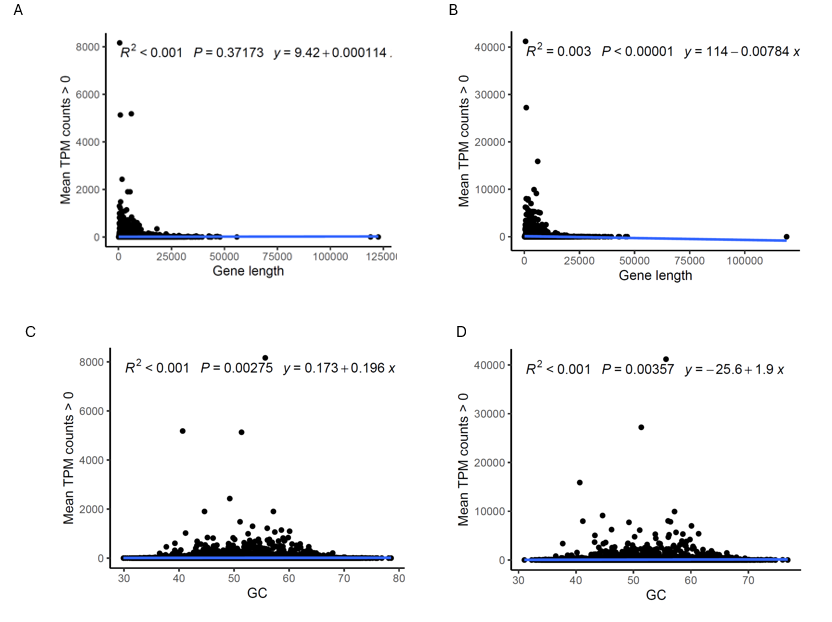

**Supplementary Figure 4. The linear regression between the length of genes and GC percentages with expression level from different platforms. A-B)** The length of genes with TPM counts from **A)** Illumina cDNA-seq from *Kallisto* and **B)** Nanopore RNA-seq from *NanoCount*. **C-D)** The GC percentages with TPM counts from **C)** Illumina cDNA-seq from *Kallisto* and **D)** Nanopore RNA-seq from *NanoCount*.

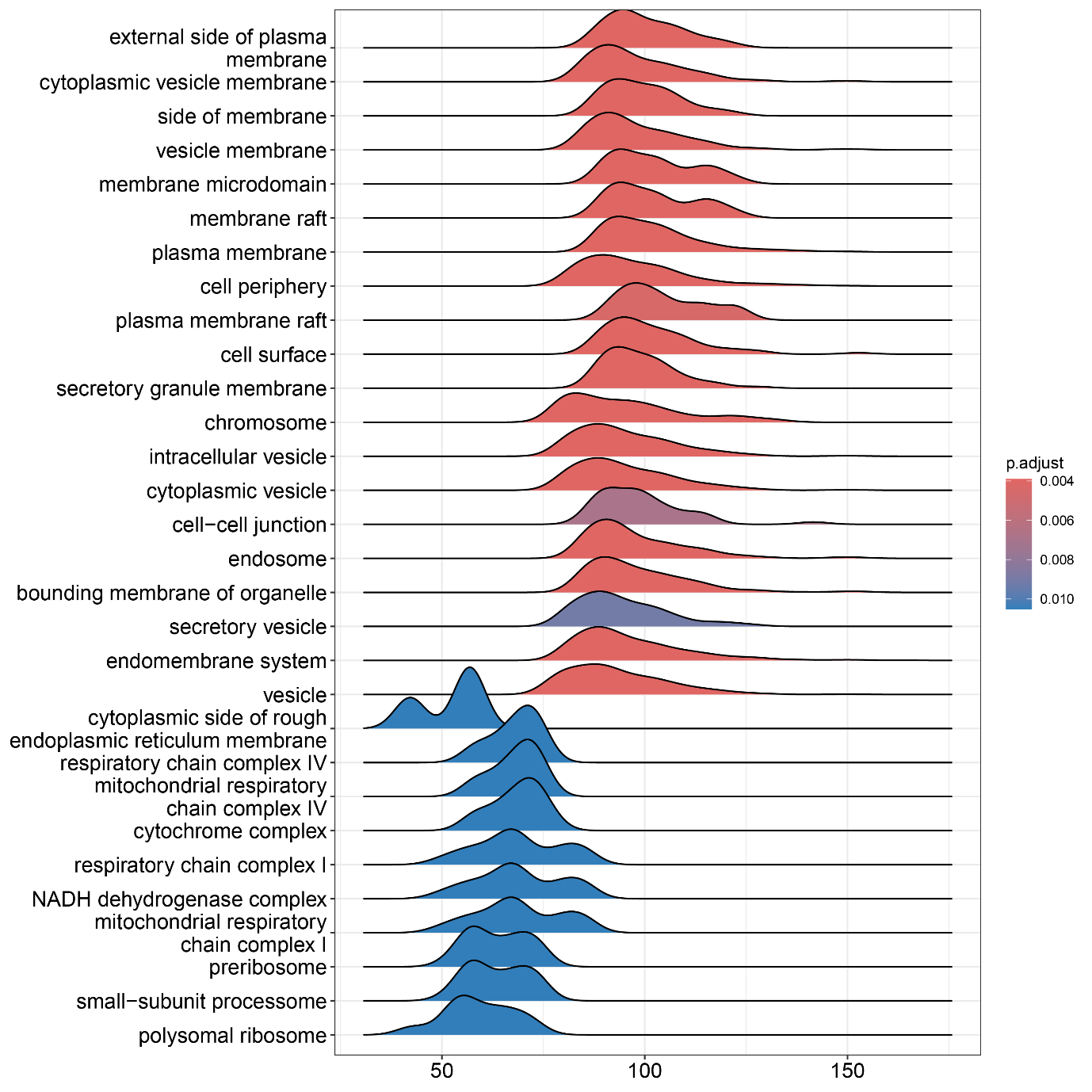

**Supplementary Figure 5.** **Ridgeplot showing the distribution of poly(A) lengths of genes that enriched in the corresponding GO term pathways (Cellular Component)**. The colour indicates the significance, with a threshold of adjusted P-value < 0.05. The X-axis indicate the Poly(A) lengths in nt and the Y-axis shows the GO terms. The full list of significant pathways can be viewed in **Supplementary Table 3**. The mitochondrial transcripts are excluded.

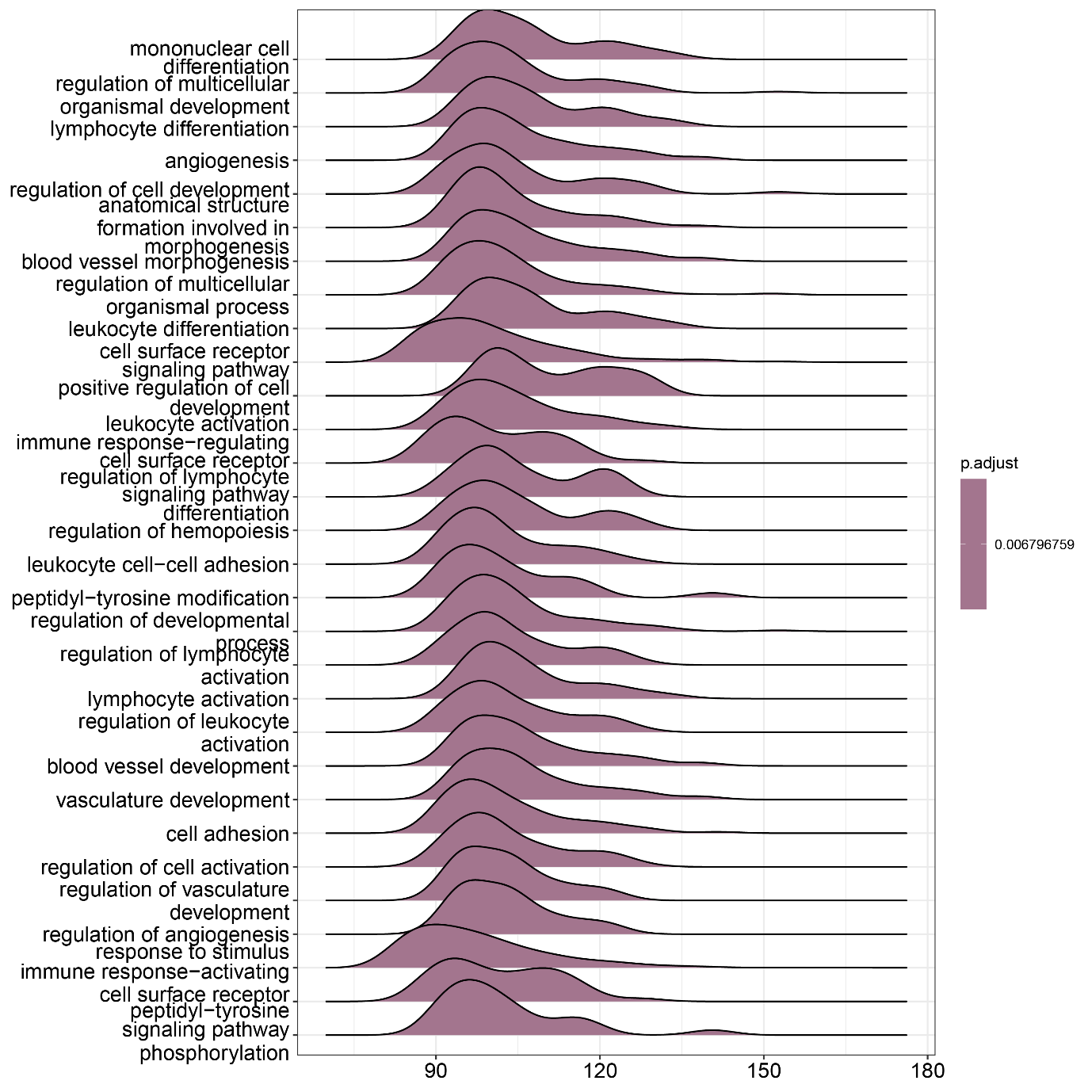

**Supplementary Figure 6.** **Ridgeplot showing the distribution of poly(A) lengths of genes that enriched in the corresponding GO term pathways (Biological Process)**. The colour indicates the significance, with a threshold of adjusted P-value < 0.05. The X-axis indicate the Poly(A) lengths in nt and the Y-axis shows the GO terms. The full list of significant pathways can be viewed in **Supplementary Table 4**. The mitochondrial transcripts are excluded.

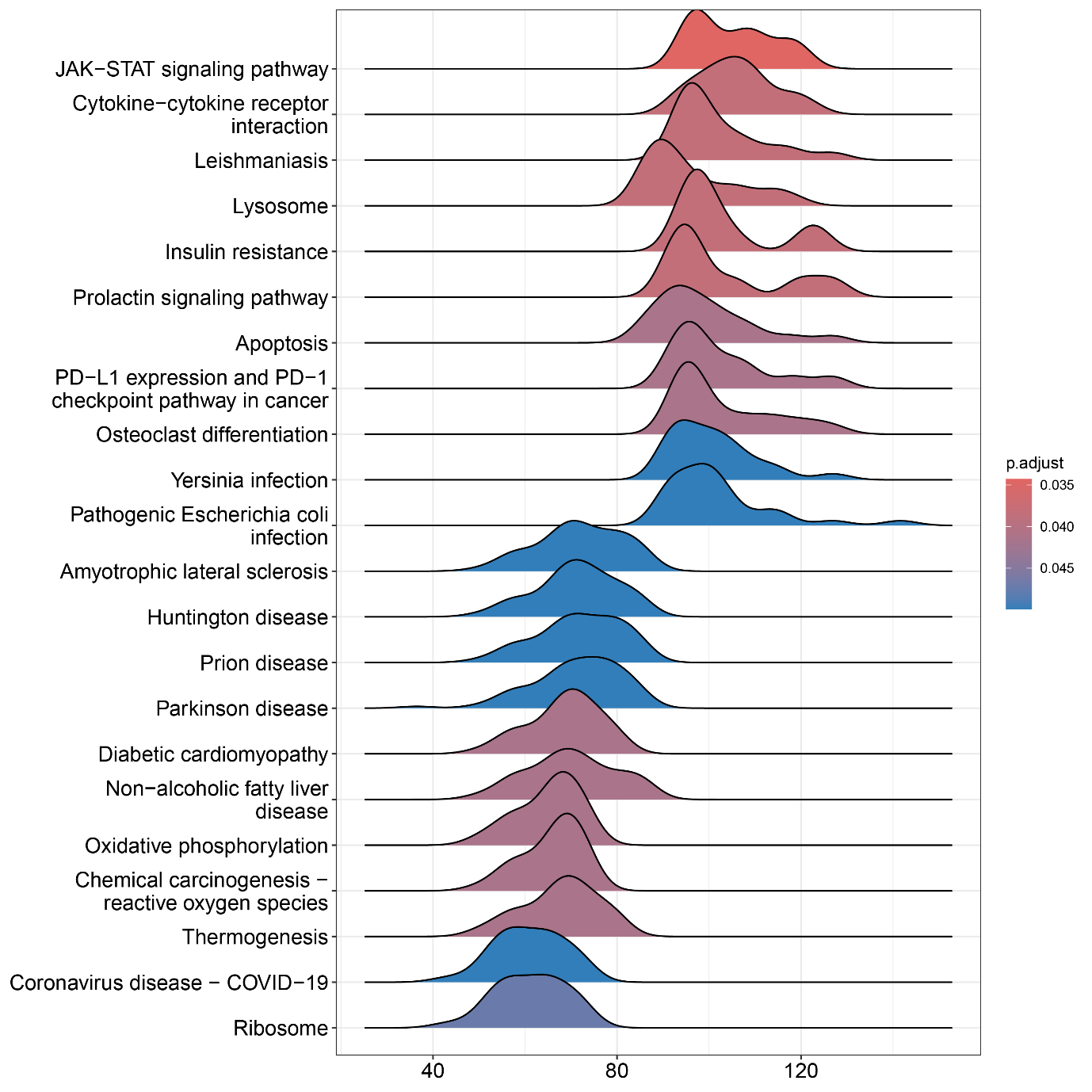

**Supplementary Figure 7.** **Ridgeplot showing the distribution of poly(A) lengths of genes that enriched in the KEGG pathways**. The colour indicates the significance, with a threshold of adjusted P-value < 0.05. The X-axis indicate the Poly(A) lengths in nt and the Y-axis shows the KEGG pathways. The full list of significant pathways can be viewed in **Supplementary Table 5.** The mitochondrial transcripts are excluded.

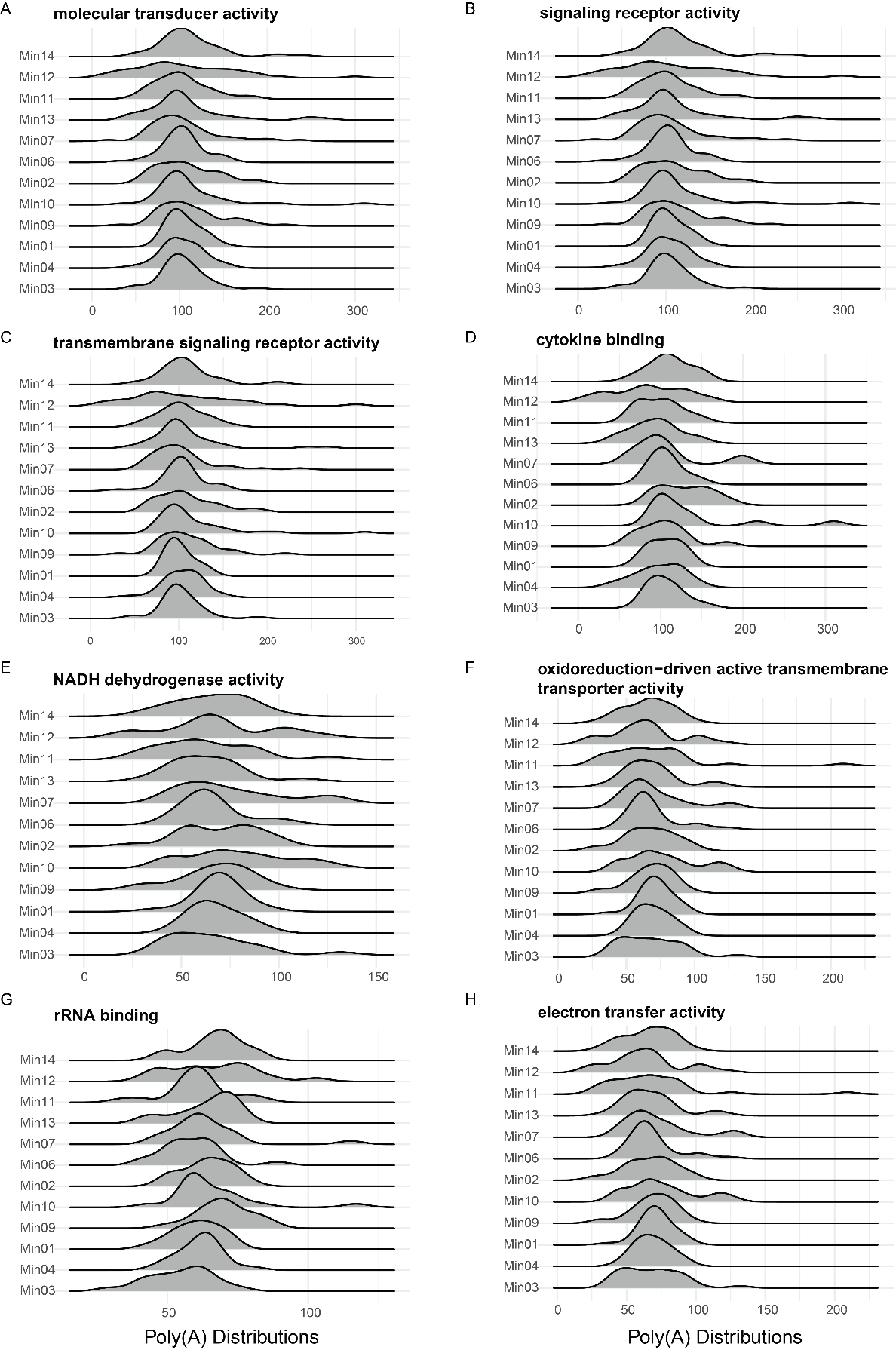

**Supplementary Figure 8.** **The poly(A) distributions for some top molecular functional pathways across 12 different samples. A-D)** The top molecular functional pathways enriched with relatively longer poly(A) tails. **E-H)** The top molecular functional pathways enriched with relatively shorter poly(A) tails. Each plot is labeled with the name of the corresponding molecular functional pathway. The X-axis indicates the poly(A) length distributions in nt and the Y-axis shows the individual data of the 12 samples.

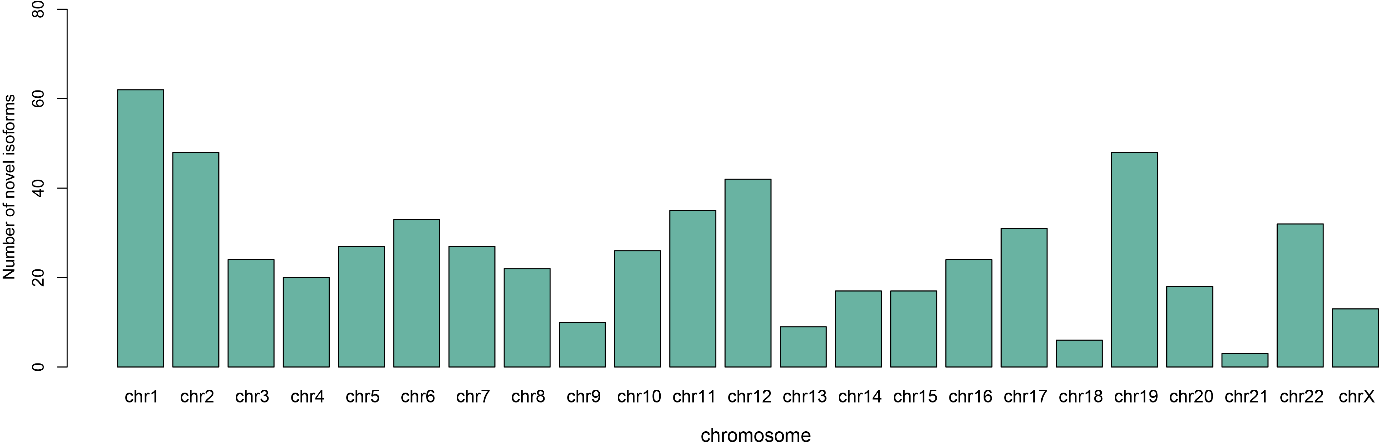

**Supplementary Figure 9. Number of novel isoforms identified by *IsoQuant* and *SQANTI3* per chromosome.**

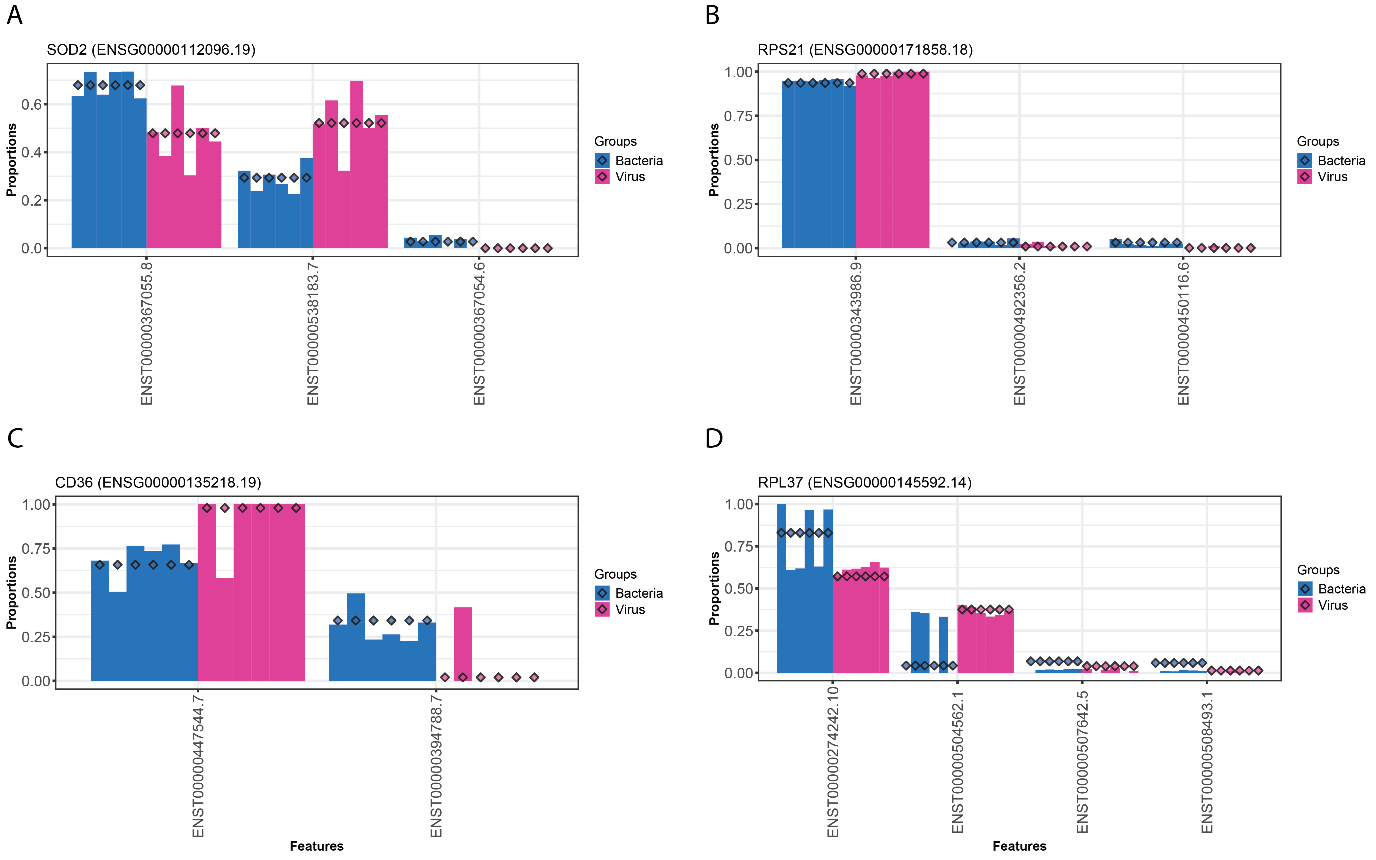

**Supplementary Figure 10. Differential transcript usage occurs between bacterial and viral samples.** **A-D)** Differential estimated proportions of transcripts of genes for **A)** *SOD2 (ENSG00000112096.19)*, **B)** *RPS21(ENSG00000171858.18)*, **C)** *CD36 (ENSG00000135218.19)*, and **D)** *RPL37 (ENSG00000145592.14)*, with adjusted P-values < 0.05.

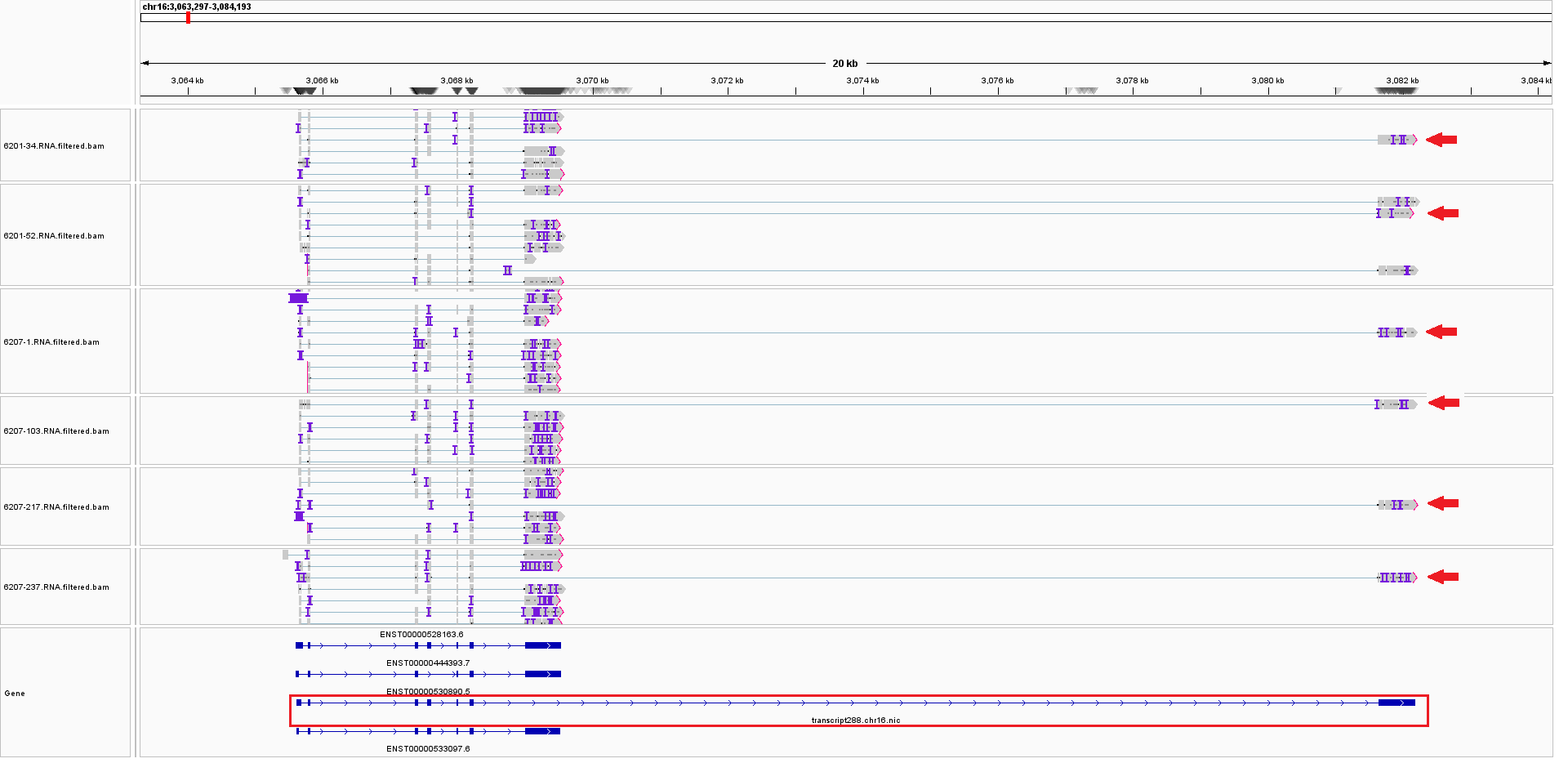

**Supplementary Figure 11. View of *IL32 (ENSG00000008517.18)* from the Integrative Genomics Viewer (IGV).** IGV-Sashimi plots showing the read coverage and transcript isoforms. The red box indicates the novel transcript *transcript 288.chr16.nic*.

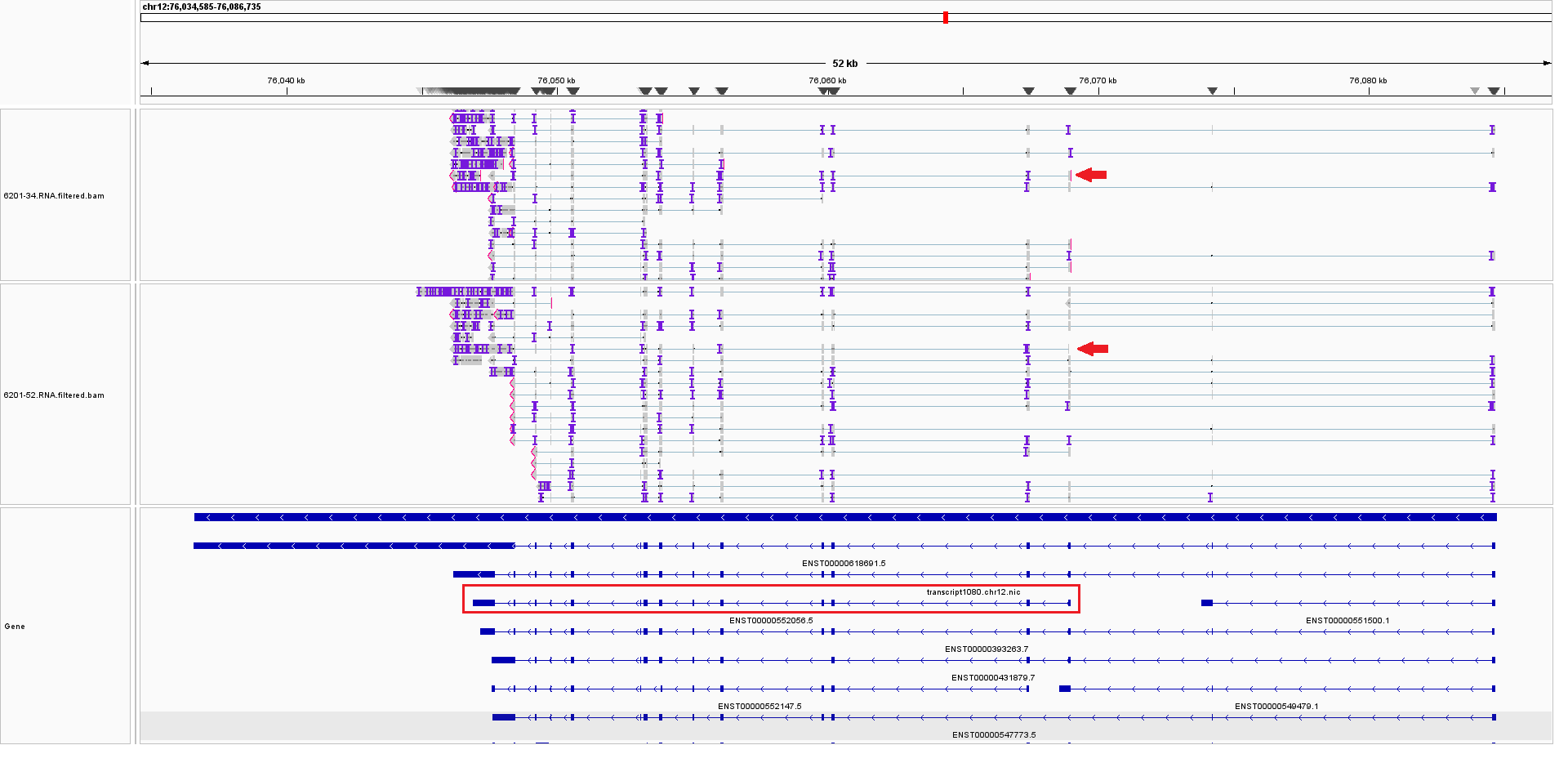

**Supplementary Figure 12. View of *NAP1L1 (ENSG00000187109.15)* from the Integrative Genomics Viewer (IGV).** IGV-Sashimi plots showing the read coverage and transcript isoforms. The red box indicates the novel transcript *transcript 1080.chr12.nic*.
